# Network-based proactive contact tracing: A pre-emptive, degree-based alerting framework for privacy-preserving COVID-19 apps

**DOI:** 10.1101/2025.08.01.25332740

**Authors:** Diaoulé Diallo, Tobias Hecking

## Abstract

Most COVID-19 exposure-notification apps still use binary contact tracing (BCT): once a test is positive, every contact whose accumulated risk exceeds a fixed threshold receives the same quarantine order. Because those alerts are late and blunt, BCT can miss early spread while triggering mass isolation. We propose Network-based Proactive Contact Tracing (NPCT), a privacy-preserving, fully decentralized intervention scheme that can run on existing exposure-notification infrastructure. Each user’s recent Bluetooth contact history is condensed into an individual risk score and compared against a dynamic, epidemic-aware threshold controlled by a single global sensitivity parameter. Crossing that threshold triggers a graded “reduce contacts by *X*%” prompt rather than an all-or-nothing quarantine. Simulations on four synthetic and empirical temporal networks show that NPCT can cut the epidemic peak by *≈* 40% while suppressing only 20% of contacts. The intervention burden concentrates on the highest-risk individuals, and the scheme’s qualitative behavior remains stable across network types, horizons and compliance levels. These properties make NPCT a practical upgrade path for national BCT apps, balancing epidemic control with privacy protection and social cost.

**Author summary:** During the COVID-19 pandemic, many countries adopted smartphone exposure-notification apps. These tools follow a binary rule: if a user’s cumulative exposure passes a fixed threshold, everyone involved is told to self-isolate. We noted two drawbacks—warnings come only after a positive test, and they can confine large numbers of people who pose little actual risk. To address this, we devised

Network-based Proactive Contact Tracing (NPCT), a fully decentralized scheme that fits inside the privacy guard-rails of existing apps. Each phone converts its owner’s recent Bluetooth encounters into a single risk score and compares that score with a threshold that tightens when case numbers rise and relaxes when they fall. Crossing the threshold triggers a request to trim only a chosen share of forthcoming contacts (for example, 25%) instead of imposing a blanket quarantine. We assessed NPCT through epidemic simulations on several synthetic and empirical temporal contact networks. The results show that this type of intervention can reduce the epidemic peak by roughly 40% percent while removing only one fifth of social interactions. NPCT therefore offers a realistic, privacy-preserving upgrade path for national exposure-notification systems.

## Introduction

Smartphone-based *digital contact tracing* (DCT) aims to warn infectious people early enough to block new clusters, flattening the epidemic curve without blanket lockdowns. Yet, apps built on the Google/Apple Exposure Notification (GAEN) framework [1] act purely *reactively*: after a positive test, a user uploads keys and any contact exceeding a fixed risk threshold receives a one-off quarantine alert. Deployments in England and Wales [2], Spain [3], and U.S. states [4] show that binary contact tracing (BCT) can curb spread at moderate uptake, but two flaws remain. First, because alerts depend on laboratory confirmation, they often arrive after secondary infections have already occurred. Second, one-size-fits-all quarantine recommendations can trigger mass isolation of low-risk contacts, undermining compliance and imposing heavy social costs.

These limitations highlight the need for *proactive*, on-device measures that continuously estimate individual risk and layer targeted interventions on top of existing BCT systems, rather than replacing them outright. Such additive strategies can issue graded, early warnings that further reduce peak prevalence and flatten the curve more effectively.

Recent work calls for *proactive* DCT that continuously updates on-device risk scores and adapts guidance to epidemic conditions [5–7]. Most schemes rely on symptom reports, centralized network visibility, or multi-bit broadcasts—choices that weaken privacy and complicate GAEN integration. Parallel studies advocate *dynamic* thresholds that tighten or loosen interventions in step with leading indicators such as infection acceleration and susceptible–infectious potential [8–11]. Yet no existing approach combines privacy-preserving risk estimation with a dynamic intervention rule responsive to real-time epidemic pressure.

We address these challenges with Network-based Proactive Contact Tracing (NPCT), a conceptual, lightweight, GAEN-compatible early-warning layer that continuously scores users by their recent Bluetooth-inferred contact activity, then applies a threshold that automatically tightens or relaxes in response to real-time epidemic indicators (infection acceleration and susceptible–infectious potential). Rather than a single blanket quarantine, NPCT issues graded “reduce contacts by *X*%” advisories—removing only a tunable fraction of each alerted user’s future interactions—and does so entirely on-device with the same privacy guarantees and cryptographic routines already deployed in national apps such as Germany’s “Corona-Warn-App”.

We evaluate NPCT by embedding it in a discrete-time SIR simulator on four temporal contact networks: three 10-day networks (two synthetic, one empirical) and one synthetic 30-day network. The most efficient configuration cuts the epidemic peak by *≈* 40% while suppressing only *≈* 20% of contacts—a two-for-one return on social cost. Across datasets, NPCT reveals a clear efficiency gradient: modest interventions yield the highest benefit per contact removed, whereas deeper peak reductions demand disproportionately greater contact suppression. These results underscore that the more aggressively one flattens the epidemic curve, the higher the associated social cost.

The remainder of the paper is organized as follows. In the Related Work section we survey existing digital contact-tracing paradigms. The Methods section details our simulation framework, risk model, dynamic thresholding rule, intervention logic, privacy considerations, and evaluation metrics. The Results section presents our empirical findings on epidemic dynamics, cost–benefit trade-offs, distributional effects, compliance sensitivity, fairness, and long-horizon robustness. Finally, Conclusion section discusses deployment implications and concludes the paper.

## Related Work

Prior research on digital contact tracing can be classified into three main paradigms: *binary* (or forward) contact tracing [7, 12, 13], which identifies and quarantines direct contacts of an index case after a positive test; *backward* contact tracing [14–16], which traces the source of infection and sibling cases in superspreading events; and *proactive* contact tracing, which continuously evaluates individual risk levels to issue early warnings based on non-binary indicators such as symptoms or contact graph proximity [5, 6, 17]. In the following, we briefly review binary contact tracing as the most widely deployed approach, before focusing in more detail on proactive contact tracing frameworks.

### Binary contact tracing

Early COVID-19 apps using the Google/Apple Exposure Notification API [1] implemented *binary* contact tracing: when a user uploads positive diagnosis keys, any contact whose accumulated exposure score exceeds a fixed threshold receives a quarantine recommendation [12, 13]. For example, the NHS COVID-19 app multiplies time-since-onset weights, indoor/outdoor context factors, distance attenuation, infectiousness kernels, and contact duration to compute per-encounter risk, summing these to decide on a single alert [18]. Data-driven variants learn these parameters from simulated exposures to improve precision [19], and fuzzy-logic extensions assign multi-level risk labels (Low/Medium/High/Too-High) for tiered advice [20]. Field studies in Spain [3], England and Wales [2], and Washington State [4] confirm that BCT can reduce transmission at moderate uptake but also drive high precautionary quarantine rates—the “pingdemic”—due to its purely reactive, one-size-fits-all alerts [21].

### Proactive contact tracing

Proactive contact tracing (PCT) continuously estimates each user’s potential to contribute to future transmission and delivers early, graded behavioral recommendations [5–7]. PCT leverages on-device features—such as recent contact volume or symptom reports—to identify users with elevated epidemiological risk. This enables tiered interventions (e.g., from increased distancing to temporary self-isolation) that reduce collective transmission risk while minimizing unnecessary restrictions on low-risk users.

Several “meta-graph” studies assume full, time-resolved network visibility and derive theoretically optimal node or edge removals, but such centralized data are incompatible with privacy-preserving deployments [22–24].

Instead, prior studies developed privacy-preserving algorithms suitable for on-device environments that continuously infer each user’s individual risk tier and deliver real-time, graded behavioral guidance. [5] demonstrated one such approach by training deep set-transformer networks on COVIsim agent-based outputs. Their smartphone-resident model predicts, for each of the previous fourteen days, an individual’s latent infectiousness using self-reported symptoms, comorbidities, Bluetooth encounters, and incoming graded risk messages; discretized risk levels are then anonymously broadcast to recent contacts. [6] subsequently proposed a rule-based extension (Rule-PCT) whose decision logic can be audited by public health authorities. Rule-PCT uses hand-crafted scoring on symptoms, pre-existing conditions, and risk messages to generate 4-level infectiousness tiers. [17] introduced a network-based data-assimilation framework that fuses proximity, test, and symptom data on a centralized dynamic graph to compute individual infection probabilities and trigger targeted isolation. A real-world implementation is Safer-Covid [25, 26], which combines on-device user factors (age, comorbidities), local incidence, and planned activity context (e.g., indoor vs. outdoor gatherings) to estimate scenario-specific risk. China’s Health Code System [27] offers maximal early intervention by mining travel history, location check-ins, and payment records to assign every citizen a color-coded status (green/yellow/red)—but does so via centralized surveillance and opaque, state-controlled algorithms [28].

### Our Contribution

Existing digital contact-tracing paradigms either remain strictly *reactive* or rely on rich personal data and centralized graph visibility that are hard to reconcile with privacy-preserving, decentralized deployments. We contribute a **Network-based Proactive Contact Tracing (NPCT)** framework that closes these gaps *and* empirically maps the cost–benefit space of such interventions.

- **Lightweight, privacy-preserving design**. NPCT infers risk solely from a user’s recent Bluetooth contact volume (temporal degree) and public case counts, requiring no symptoms, demographics, location trails or multi-bit exchanges—making it drop-in compatible with GAEN-style apps.
- **Epidemic-aware adaptivity**. A single dynamic threshold—driven by infection acceleration and susceptible–infectious potential—tightens or relaxes guidance in real time, enabling earlier but selective alerts without manual retuning as incidence changes.
- **Quantified cost–benefit envelope**. Across four temporal networks we show that modest, selective interventions can cut the epidemic peak by ≈ 40% while suppressing only ≈ 20% of contacts, and we chart the diminishing-returns frontier where deeper peak reductions demand disproportionately larger social cost.
- **Fairness and robustness**. NPCT directs most contact suppression toward the highest-risk users, maintains a proportionate burden as intervention sensitivity or strength changes, and preserves much of its behavior even with partial compliance and across month-long horizons.

Together, these findings demonstrate that NPCT not only adds a privacy-preserving proactive layer on top of BCT but also delivers a favorable, well-characterized efficiency profile that can inform public health calibration of digital interventions.

## Methods

We introduce a network-based proactive digital contact-tracing (NPCT) framework that tailors interventions to each user’s recent contact patterns and the epidemic’s current trajectory. This section explains our simulation pipeline—temporal networks, discrete-time epidemic model, risk scoring, dynamic thresholding, and the resulting intervention logic—and closes with privacy considerations that permit fully decentralized deployment using only on-device data.

### Simulation setup

We evaluate our framework by simulating disease spread and adaptive interventions on temporal contact networks using a discrete-time susceptible–infectious–recovered (SIR) model. Each simulation proceeds over a series of hourly snapshots, with the contact network changing over time. At the core of our approach is an online intervention pipeline: at regular intervals, we evaluate recent contacts, assign risk scores, update a global risk threshold based on epidemic indicators, and apply selective contact suppression to high-risk individuals. This cycle mimics the on-device NPCT loop, including real-time, graded contact reduction prompts. Between these intervention points, the disease propagates naturally on the evolving network.

More precisely, interventions are applied at discrete times *T*_0_, *T*_1_, …, spaced by an interval Δ*t*. At each *T*_*j*_, we evaluate the current state to assess recent contacts, compute risk scores, update the intervention threshold, and suppress a fraction of future contacts for high-risk individuals; the cycle repeats until simulation end. Full details of the risk computation, thresholding mechanism, and intervention design appear in the Risk modeling, Dynamic intervention threshold and Intervention mechanism sections.

### SIR model

We simulate a discrete-time SIR process on each temporal network. At *t* = 0 we randomly seed 5% of nodes as infectious and leave the remaining 95% susceptible. In each subsequent snapshot, every infectious node attempts to infect each of its susceptible neighbors with transmission probability *β*, and independently recovers with probability *γ*.

Because temporal networks lack a closed-form epidemic threshold [29], we conducted a brief parameter sweep (S2 Appendix) to identify transmission and recovery rates that produce a pronounced epidemic curve with a well-defined infection peak. The selected *β* and *γ* values are displayed in Table 1. These parameter choices yield a pronounced epidemic peak, giving headroom to measure intervention effects.

**Table 1.** Network size, duration, and SIR parameters for each dataset.

| Network | #Nodes | #Edges | #Timesteps | $\beta$ | $\gamma$ |
| --- | --- | --- | --- | --- | --- |
| ABM30 | 2058 | 159173 | 720 | 0.005 | 0.002 |
| ABM | 2058 | 67778 | 240 | 0.010 | 0.005 |
| DTU | 691 | 42453 | 240 | 0.040 | 0.005 |
| Office | 92 | 681 | 240 | 0.300 | 0.005 |

We compare each scenario with and without NPCT on an identical SIR backbone. Every parameter configuration runs 200 stochastic realizations produced by a shared set of 200 random seeds, ensuring one-to-one comparability. Extensive network-epidemiology studies show that it is the contact network—rather than the presence of an exposed (latent) stage—that determines both the epidemic peak and final size; adding a latency period simply delays and uniformly stretches the corresponding SIR outbreak curve without changing its shape [30–32]. Accordingly, an SIR core suffices to reveal NPCT’s intrinsic cost–benefit profile on realistic temporal networks; richer disease-state extensions can be layered in future work. We believe our simulation experiments thus make a valuable contribution to understanding how NPCT can flatten the curve under realistic contact patterns.

### Datasets

#### ABM network

For our experiments, we use the publicly available TCN1000-medium temporal contact network introduced in our previous work [33, 34]. We refer to this as the *ABM* network. It comprises 2,058 individuals organized into 1,000 households and spans five location types. The network covers a 10-day period at hourly resolution, resulting in 240 temporal snapshots, generated by the MEmilio–HuMM framework and calibrated against high-resolution empirical references. Full details of its generation methodology, capacity settings, and mobility model calibration can be found in the original publication [33]. It is particularly well-suited for evaluating digital contact tracing, as it represents a small but demographically diverse population (multiple age groups and household compositions) interacting across a variety of empirically calibrated location types and sizes—households, schools, workplaces, supermarkets, social events—and exhibits realistic temporal patterns (day–night cycles, bursty contacts, heavy-tailed degree distributions) that mirror real-world mobility and social mixing.

#### ABM30 network

To assess NPCT over a longer timeframe, we extend the ABM network to 30 days (*ABM30*) by running the same MEmilio–HuMM generation pipeline. This 30-day synthetic network preserves the demographic diversity, location types, and realistic contact dynamics of the original 10-day ABM dataset, enabling evaluation of NPCT’s temporal robustness.

#### DTU network

The *DTU* temporal contact network is drawn from the Copenhagen Networks Study [35], including 691 students from the Technical University of Denmark. Via study-issued smartphones, the dataset provides a multi-layer temporal network—Bluetooth-inferred physical proximity, phone calls, SMS, and Facebook friendships—all at high temporal resolution. Here we make use of the Bluetooth proximity layer, aggregated to an hourly time step over a 10-day window, yielding 240 snapshots. As one of the few publicly available human contact datasets spanning at least ten days with fine-grained, real-world proximity measurements, the DTU network offers a valuable benchmark for evaluating digital contact tracing methods.

#### Office network

The *Office* temporal contact network captures face-to-face proximity among employees of a French public health institute [36]. During a two-week period, volunteers agreed to wear ultra-low-power RFID badges that record any face-to-face encounter lasting at least 20 seconds, with a spatial resolution of approximately 1.5m and a temporal resolution of 20s. For consistency with our other datasets, we aggregate the raw proximity events into hourly snapshots and restrict the analysis to the first ten days of the collection, yielding 240 temporal layers. The resulting temporal network comprises 92 active nodes. This compact, empirically grounded benchmark offers a realistic work environment for evaluating proactive contact tracing interventions.

### Risk modeling

Let the temporal contact network be represented as a sequence of snapshots

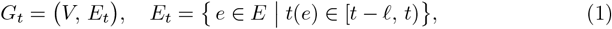

where *V* is the fixed node set, *E* the set of all time-stamped edges, *t*(*e*) the timestamp of edge *e*, and *𝓁* the snapshot window length of one hour.

Also, let *k*_*i*_(*t*) denote the instantaneous degree of node *i* in snapshot *G*_*t*_. We quantify the *temporal degree*—used here as a risk proxy—of node *i* over the latest intervention window Δ*t* as

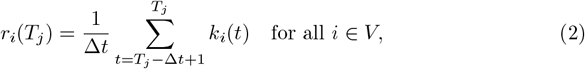

where *T*_*j*_ is the current intervention time (see Fig 1). To ensure comparability across nodes and timesteps, and to improve the dynamic range of values in [0, 1], we apply a logarithmic normalization to the raw risk scores.

**Fig 1.**
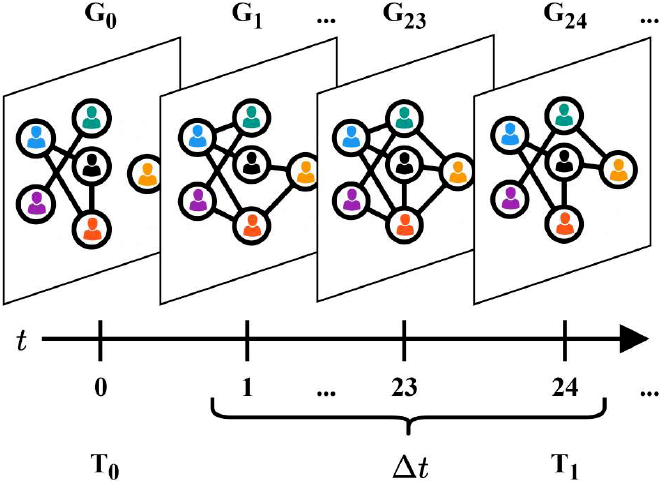
Temporal network snapshots. The first intervention is at *T*_1_ = 24, given that Δ*t* = 24.

Let *r*_max_ denote the global maximum of all observed degrees across all nodes and all timesteps of *G*_*t*_:

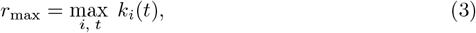

In practical deployment, this value can be approximated beforehand using historical contact data.

We define the normalized risk score as

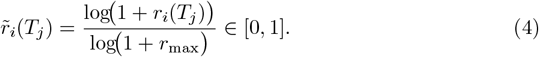

Log–scaling preserves node ranking while preventing extreme degrees from dominating, distributing scores more evenly over [0, 1] and simplifying thresholding.

Although more sophisticated metrics can identify vital, high-impact nodes more precisely, they typically require more network visibility; our earlier study showed that simple degree retains much of their predictive power while using only the immediate neighborhood of a node, thus respecting stricter privacy constraints [37].

### Dynamic intervention threshold

#### Epidemic acceleration

Let *I*(*t*) be the number of infectious individuals at step *t*. Given a look-back window *w*, we define the (clamped) infection acceleration at intervention time *T*_*j*_ as

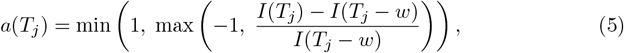

which lies in the interval *a* ∈ [−1, 1]. Here, *a* > 0 indicates a growing epidemic, while *a* < 0 signals a decline in the number of active infections [38–40]. This clamping improves stability by avoiding extreme threshold shifts.

#### Global infection potential

Drawing on the mass-action interaction term *SI* from classical compartmental models, we define a dimensionless global infection potential:

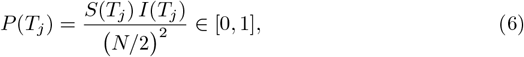

where *S*(*t*) and *I*(*t*) denote the number of susceptible and infectious individuals at time *t*, and *N* = |*V* | is the total population size. We normalize *S* · *I* by its theoretical maximum (*N/*2)^2^, reached when half of the population is susceptible and half is infectious. As a result, *P* (*T*_*j*_) reaches its maximum value of 1 when the population is most epidemiologically “active,” and drops to 0 when either compartment dominates. The potential thus captures the current number of potentially infectious interactions relative to the most transmissive configuration possible.

#### Threshold update mechanism

At each intervention time *T*_*j*_, the algorithm maintains a risk threshold *θ*(*T*_*j*_) ∈ [0, 1], which determines which nodes are considered high-risk. A node *i* is selected for intervention if its normalized risk score 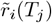 exceeds this threshold. The initial value is set to *θ*(*T*_0_) = 1, and is updated thereafter based on two epidemic indicators: infection acceleration and global infection potential.

To form a composite signal reflecting current epidemic pressure, we compute a weighted combination of the clamped infection acceleration *a*(*T*_*j*_) and infection potential *P* (*T*_*j*_):

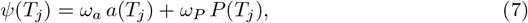

where *ω*_*a*_, *ω*_*P*_ > 0 are fixed weighting parameters. In our experiments, both are set to 1.0, assigning equal influence to acceleration and infection potential.

The two components of *ψ*(*T*_*j*_) offer complementary dynamics: the global infection potential *P* (*T*_*j*_) is structurally smooth, as it depends on the aggregate counts of susceptible and infectious individuals. Since these quantities typically evolve gradually, *P* (*T*_*j*_) changes slowly and anchors the threshold update in the longer-term transmission landscape. In contrast, the infection acceleration *a*(*T*_*j*_) reflects short-term changes in epidemic growth. Because it captures the relative increase or decrease in active infections over a recent window, it introduces a form of temporal memory and enables the system to react more swiftly to emerging trends. Their combination ensures that the threshold remains both responsive and stable across different phases of the epidemic.

The composite value *ψ*(*T*_*j*_) is then mapped to a final target threshold via a linear sensitivity rule,

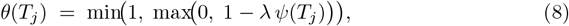

where *λ* > 0 scales the influence of the epidemic pressure on the threshold, starting each time from the baseline value 1. Because *a*(*T*_*j*_) ∈ [−1, 1] and *P* (*T*_*j*_) ∈ [0, 1], the composite signal is bounded by *ψ*(*T*_*j*_) ∈ [−1, 2]. In practice, the lower bound −1 would require a simultaneous zero infection potential and maximal negative acceleration—an unlikely combination—so typical values lie in a narrower range. The clamping ensures that the threshold can fall by at most 2*λ* and rise by at most *λ*. This built-in asymmetry implements a precautionary principle: the threshold can drop quickly when pressure rises, but it relaxes more conservatively as the epidemic recedes. We vary *λ* in our experiments to explore its impact on intervention dynamics. This design balances interpretability while offering flexibility: it allows for responsive threshold adaptation to both rising and falling epidemic trends, with *λ* tuning sensitivity.

### Intervention mechanism

Interventions are applied at discrete times *T*_0_, *T*_1_, …, each spaced by Δ*t* = 24 hours, so that *T*_*j*_ occurs once per day.

At time *T*_*j*_ we construct the high-risk set

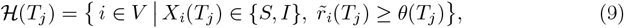

where *X*_*i*_(*t*) ∈ *{S, I, R}* is node *i*’s epidemic state. For every *i* ∈ ℋ(*T*_*j*_), we randomly delete a fraction *ϕ* ∈ [0, 1] of its edges in future snapshots *G*_*t*_ for *t* ∈ (*T*_*j*_, *T*_*j*_ + Δ*t*].

When *ϕ* = 1 the procedure mimics strict quarantine; 0 < *ϕ* < 1 models partial behavioral change such as limiting social contacts. This mechanism mimics behavioral changes induced by digital exposure notifications, as could be implemented in decentralized contact tracing (DCT) apps, where users are advised to reduce contacts upon receiving a risk alert. Random pruning overlooks that users typically retain core contacts (e.g., household edges) and prune only discretionary ties; however, under the simplifying assumption of a uniform per-contact transmission probability *β* and hourly-resolution snapshots, removing a random *ϕ*-fraction of edges approximates the expected reduction in exposure and thus provides a conservative, lower-bound estimate of NPCT’s impact. Future work should incorporate contact-type–specific pruning rules to better reflect real behavioral choices.

To reflect the fact that not all users comply with such advice, we introduce a *compliance parameter c* ∈ [0, 1], which scales the intended removal fraction. The effective fraction of contacts removed is thus *c ϕ*, where *c* = 1 denotes perfect adherence and *c* = 0 corresponds to no behavioral response. In addition to the main experiments (with *c* = 1.0), we sweep *c* ∈ {0.3, 0.6, 0.9, 1.0}. This allows us to quantify how NPCT’s epidemic-control potential—measured via peak reduction and curve flattening—depends on compliance.

### Privacy considerations

NPCT is designed to work without ever exporting personal contact data. Each phone computes its own temporal-degree risk score from locally stored Bluetooth histories; the scalar value 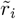 stays on the device and is compared only to a global threshold *θ*. The adaptive threshold uses epidemic acceleration and infection-potential indicators derived from anonymized, aggregated case counts—no individual graph data or health states are required. Interventions are triggered only for users whose local score exceeds *θ*, avoiding blanket restrictions and further limiting data exchange. Thus, NPCT achieves targeted,network-aware mitigation while preserving full on-device privacy.

### Evaluation metrics

Digital contact-tracing interventions are useful only insofar as the epidemiological benefit outweighs the social cost of disrupted contacts. We therefore report two dimensionless *efficiency ratios* :

#### Peak-efficiency ratio (ℰ_peak_)

Let *I*_baseline_(*t*) and *I*_int_(*t*) be the prevalence under baseline and a given intervention, respectively, and let

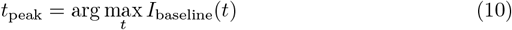

be the time of peak prevalence in the baseline. We then define the absolute peak reduction

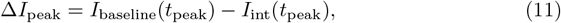

and let *C*_edges_ be the cumulative percentage of edges removed over the entire simulation. The peak-efficiency ratio is

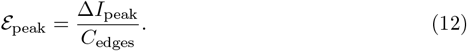

Values ℰ_peak_ > 1 indicate that each percent of edges removed yields more than a one-percent reduction in the infection peak, indicating that the intervention yields epidemiological benefit per unit of social cost.

#### Attack-rate efficiency (ℰ_AR_)

Analogously, let Δ*I*_cum_ be the absolute reduction in cumulative infections (final attack rate) and reuse *C*_edges_. Then

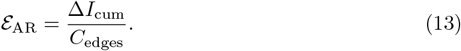

## Results

Performance is evaluated across four datasets—ABM, ABM30, DTU, and Office—by examining how NPCT configurations influence epidemic progression, network structure, and intervention efficiency. The three 10-day networks are analyzed first, tracking the co-evolution of infections, edge removals, and the adaptive threshold under varying sensitivities and intervention strengths. Next, the cost–benefit profile is quantified by comparing peak reduction and cumulative case suppression against the burden of contact suppression. This is followed by an investigation of distributional effects, assessing how risk and intervention burdens concentrate across the population.

Subsequently, the impact of decreased compliance levels is presented. For all main experiments, we assume a compliance of *c* = 1. In addition to risk and intervention distributions, we also characterize whether social costs are allocated fairly. Finally, the 30-day ABM30 network is examined to assess long-horizon dynamics, intervention characteristics, and efficiency.

NPCT is designed as an add-on layer that can run alongside the binary contact-tracing (BCT) logic already embedded in GAEN-style apps: it works entirely on device and adapts in real time to epidemic acceleration and infection potential. Classical BCT, by contrast, hinges on an external and highly variable chain—symptom-to-test delays, laboratory turnaround, key-upload timing, and country-specific risk thresholds—plus non-trivial false-positive and false-negative rates. Because these parameters differ widely across deployments and are rarely available at sufficient temporal resolution for simulation, a head-to-head comparison between NPCT and BCT would be neither robust nor generalizable. Instead, we focus on mapping NPCT’s internal cost–benefit envelope across different intervention strengths and compliance levels. Throughout this section, we benchmark NPCT against two reference points: the standard no-intervention baseline, where the epidemic unfolds on the raw temporal network; and the no-threshold baseline (*λ*→∞), in which every node is categorized as high-risk at every intervention step—effectively mirroring a blanket “reduce contacts” recommendation that ignores the current epidemic state and network heterogeneity.

As a reminder, *λ* is the global sensitivity that governs how epidemic pressure shifts the adaptive threshold (Eq. 8), *ϕ* is the intended fraction of future contacts removed for high-risk individuals (see Intervention mechanism), and *θ*(*T*_*j*_) denotes the threshold value at intervention time *T*_*j*_ (Eq. 8); a complete list of symbols appears in Table S1 Appendix.

### Epidemic and Network Dynamics

Fig 2 shows the progression of three key quantities: infections over time, edges remaining over time, and the evolution of the dynamic intervention threshold. Equivalent plots for the DTU and Office datasets are provided in S3 Appendix. The left most column of Fig 2 presents infection prevalence, with time on the x-axis (in hours) and the percentage of infectious individuals on the y-axis. The gray curve represents the baseline scenario without any intervention, while colored curves indicate NPCT simulations for varying sensitivity values *λ* ranging from 0.5 to 2. The middle column reports the percentage of edges remaining (relative to baseline) in the contact network at each time step, again plotted over time in hours and stratified by *λ*. The right most column shows how the adaptive threshold *θ*(*T*_*j*_) evolves over the course of the simulation. Here, the x-axis denotes the intervention step, ranging from 0 to 9, as the simulation applies interventions once per day over a total duration of ten days. At step 0, no intervention has yet occurred; subsequent steps show how the threshold adapts in response to the epidemic state. Each row of Fig 2 corresponds to a different removal fraction *ϕ* ∈ *{*0.10, 0.25, 0.50, 1.00*}*, where *ϕ* = 1.00 represents full quarantine of the high-risk set, and lower values represent proportionally weaker contact-reduction recommendations. Together, these plots illustrate how NPCT shapes epidemic and network behavior under different intervention strengths and sensitivity settings.

**Fig 2.**
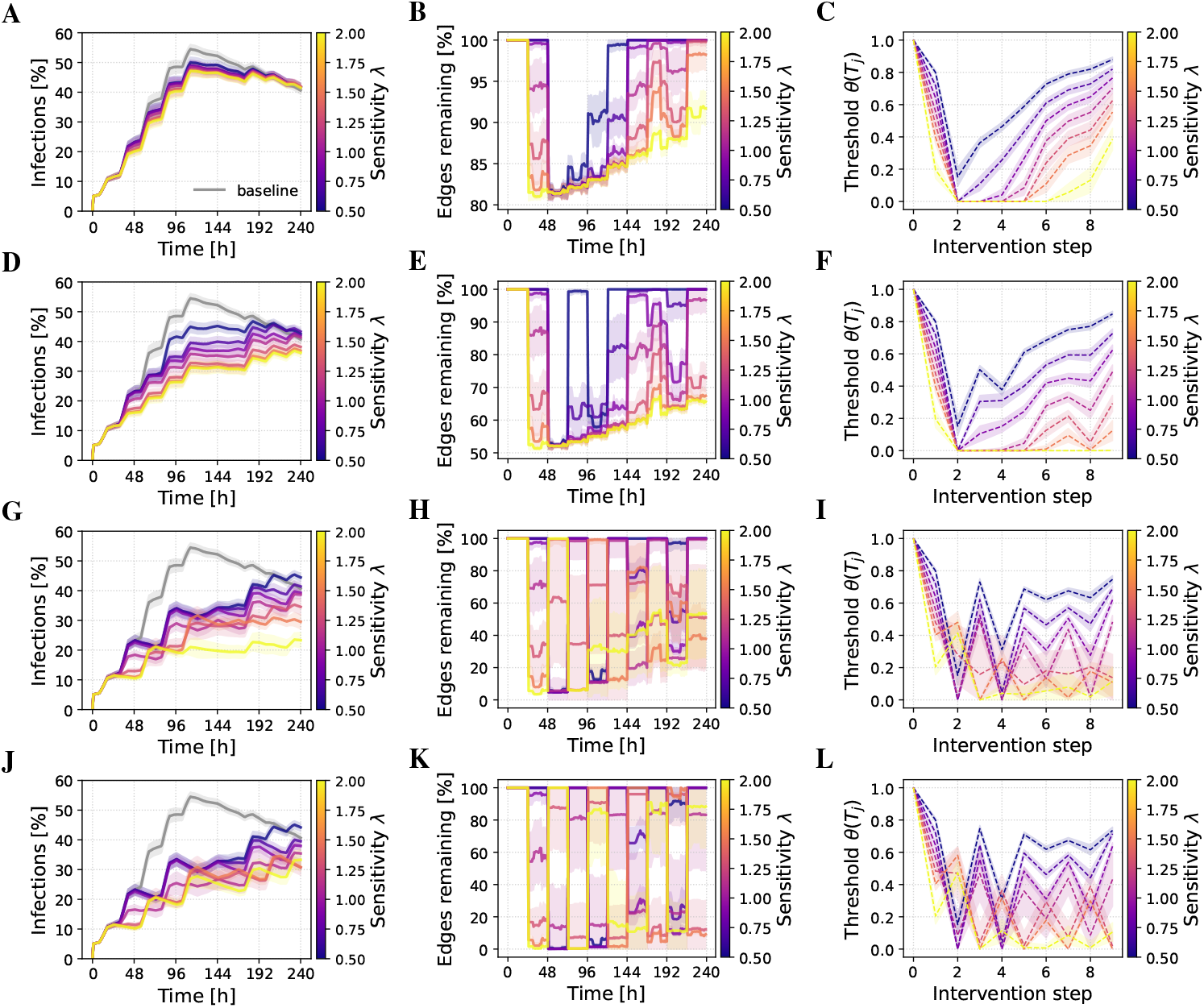
Progression of key quantities under NPCT interventions for ABM. (A, D, G, J) Infection prevalence over time (hours, x-axis; percentage infected, y-axis), with the gray curve representing the no-intervention baseline. (B, E, H, K) Percentage of edges remaining in the contact network (relative to baseline) over time (hours). (C, F, I, L) Evolution of the adaptive threshold *θ*(*T*_*j*_) across intervention steps. Colored curves represent sensitivity *λ* ∈ [0.5, 2], results are averaged over SIR runs. Each row corresponds to a removal fraction *ϕ* ∈ {0.10, 0.25, 0.50, 1.00} (top to bottom). Identical panels for the DTU and Office datasets are available in S3 Appendix.

We observe a clear dependence of epidemic control on both the sensitivity parameter *λ* and the removal fraction *ϕ*. With the weakest intervention (*ϕ* = 0.10, top row) the peak prevalence falls by roughly 5–10 percentage points, with higher *λ* values having slightly more impact. The threshold trajectories account for this difference. High sensitivities drive the cutoff from 1.0 to ≈ 0.2 after the first daily update and keep it low for several days, classifying many users as high-risk. Low sensitivities, by contrast, produce shallower dips that rebound quickly, so far fewer contacts are suppressed. The sharp drop in *θ* between the first and second interventions is driven by epidemic acceleration, which is highest at the start of the outbreak. A broader depression appears later, when the product *SI* peaks and the infection-potential term then dominates the composite signal. The middle column confirms that network pruning follows the same ordering: large *λ* values trigger edge removals earlier and sustain them longer, whereas small *λ* values delay the onset of interventions and restore connectivity soon after the threshold recovers. A modest resurgence of infections toward the end of the ten-day window stems from changes in the contact pattern; only the high-sensitivity settings respond by reinstating edge removals.

At *ϕ* = 0.25, the most aggressive setting (*λ* = 2.0) reduces the infection peak to roughly half of the baseline. For intermediate and high sensitivities (*λ* ≥ 1.00), we observe the classic “flatten-the-curve” effect: prevalence increases more slowly and never reaches the sharp maximum seen in the baseline. Notably, while the baseline scenario exhibits a clear peak around hour 110 followed by a decline, many NPCT configurations with stronger interventions display a continued rise in infections until the end of the 10-day window. This extended growth phase reflects the fact that earlier interventions leave more susceptible individuals in the population, delaying saturation effects. A quantitative comparison of peak reduction and intervention efficiency across all datasets follows in the Intervention Efficiency section.

At higher removal fractions (*ϕ* = 0.50 and *ϕ* = 1.00), we observe greater variability (standard deviation) and fluctuations in both the threshold evolution and the fraction of edges removed. This behavior stems from two interacting effects. First, strong interventions—such as cutting half or all of a user’s contacts—immediately reduce the individual’s future contact activity, and thus their temporal degree and risk score. As a result, many of the nodes targeted in one intervention step fall below the threshold in the next, causing a rapid recovery in the number of edges and producing an oscillatory pattern in the pruning curves. Second, these strong interventions also suppress the spread of infection more effectively, leading to lower acceleration and reduced epidemic pressure. This weaker composite signal, in turn, drives the threshold upward again.

These effects are clearly visible in the threshold plots: in contrast to *ϕ* = 0.10 and *ϕ* = 0.25, where thresholds mostly decline and stabilize, the curves for *ϕ* = 0.50 and *ϕ* = 1.00 often exhibit an early rebound followed by fluctuating trajectories. This oscillatory behavior reflects the on–off nature of the intervention dynamics, where strong suppression alternates with temporary relaxation, allowing new infections to emerge before the system responds again.

Beyond *ϕ* = 0.5, returns taper off: infection curves for *ϕ* = 0.5 and 1.0 look nearly identical, suggesting that beyond a certain intervention strength, further increasing *ϕ* yields limited additional benefit. In fact, for the most aggressive sensitivity setting (*λ* = 2.0), the infection curve is slightly flatter for *ϕ* = 0.50 than for *ϕ* = 1.00. Because full quarantine preserves a larger susceptible population, transmission can rebound more sharply once interventions are lifted. At *ϕ* = 0.50, the system maintains a more continuous level of contact suppression, resulting in smoother epidemic control. These findings highlight the interplay between intervention strength, temporal structure of contacts, and the evolving distribution of susceptibility within the network. Moreover, given the finite temporal horizon and the extensive overlap in contact patterns, once a critical mass of high-risk individuals has already had edges removed during one intervention step, further edge removals increasingly target the same—or now absent—contacts, leading to a natural plateau in marginal epidemic impact. This saturation behavior is also apparent in the compliance sweeps (Impact of user compliance section).

### Intervention Efficiency

#### Cost-benefit Profile of Risk-Based Interventions

Fig 3 illustrates the trade-off between epidemiological benefit and intervention cost on all four networks. In the top row, bubbles plot peak-infection reduction against the percentage of edges removed; in the bottom row, bubbles plot the same measure against the cumulative number of high-risk notifications. Bubble size represents the removal fraction *ϕ* ∈ *{*0.1, 0.2, 0.3, 0.4, 0.5, 0.6, 0.75, 1.0*}*, and color encodes the sensitivity parameter *λ*. Peak reduction is measured at the baseline’s epidemic peak, whereas both cost metrics are accumulated over the full simulation.

**Fig 3.**
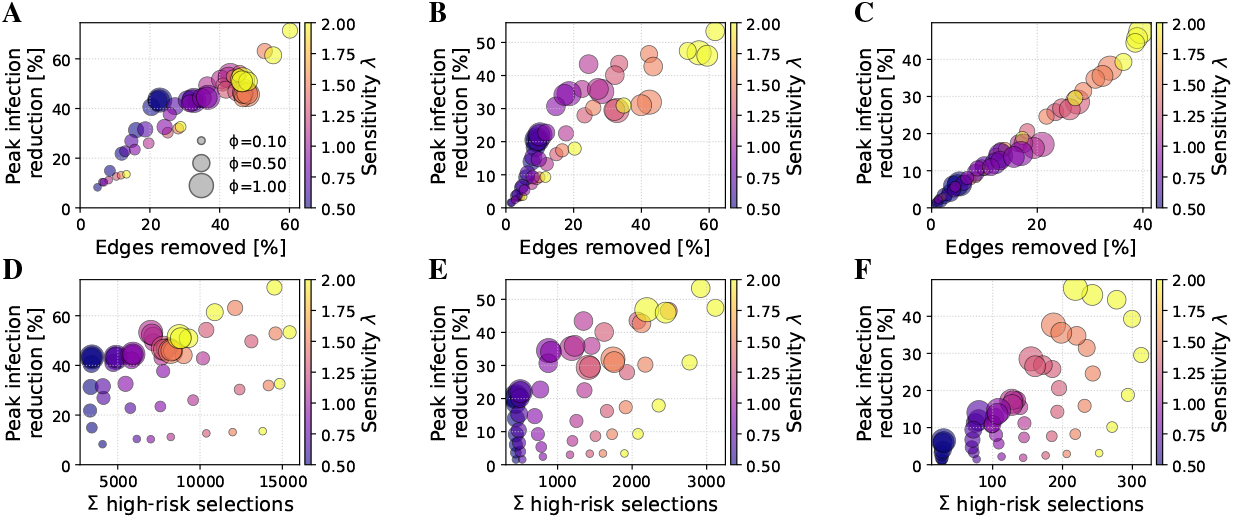
Bubble-plots of peak infection reduction vs. cost. Results are shown for ABM (A, D), DTU (B, E), and Office (C, F). The top row shows peak infection reduction versus percentage of edges removed; the bottom row shows peak infection reduction versus total high-risk notifications sent (high risk selections). Bubble area encodes removal fraction *ϕ*, color indicates sensitivity *λ*, results are averaged over SIR runs.

Focusing on the top row (A, B, C), we observe that for both the ABM and DTU networks, lower sensitivity values (*λ* ≈ 0.5) tend to result in limited intervention effects—both in terms of edges removed and peak reduction—suggesting that low sensitivity delays or suppresses response to epidemic pressure. These low-*λ* settings often achieve favorable cost-benefit tradeoffs: moderate reductions in peak prevalence are achieved with relatively few edge removals. In contrast, higher sensitivity values (*λ* > 1.0) consistently produce larger peak reductions, but at a steeper cost in network disruption. This yields diminishing returns: while absolute impact improves, the efficiency (reduction per edge removed) worsens. For instance, in the DTU case, *λ* = 1.5 produces one of the worst cost-benefit ratios, with less than 1% reduction in peak per percent of edge removal. The ABM network shows a more favorable scenario, where the most efficient configuration achieves a ≈40% peak reduction with ≈20% edge removal (efficiency factor ≈ 2).

Across both ABM and DTU, we also observe saturation effects: beyond ≈20–25% of edges removed, additional pruning yields reduced gain in peak reduction. The role of the removal fraction *ϕ* appears primarily multiplicative: it scales the intervention impact, but does not qualitatively alter the shape of the performance curve for a given *λ*. In other words, increasing *ϕ* shifts the corresponding bubbles upward and rightward, but the relative ordering of *λ*-traces remains similar. Notably, for high *λ*, the largest *ϕ* does not always produce the strongest impact—suggesting nonlinear dynamics or possible oversuppression, where early intervention lowers risk scores in subsequent rounds and weakens sustained effect.

Results on the Office data set vary far less: peak reduction scales almost linearly with edges removed, and curves lie close to a 1:1 line. This behavior reflects the network’s homogeneous, sparse structure, which offers fewer opportunities for targeted pruning to deliver outsized gains. As observed for the other networks, *ϕ* has the greatest effect at low *λ*, with its influence saturating as sensitivity increases.

Beyond edge removals, intervention cost also depends on how many users are targeted. The bottom row of Fig 3 plots peak reduction against the total number of high-risk selections. A clear trend emerges: increasing the sensitivity parameter *λ* yields more frequent or broader targeting, with cumulative selections rising approximately linearly with *λ* across all datasets. This reflects the stronger threshold response to epidemic pressure at higher sensitivities, leading to more individuals being classified as high-risk over time. The Office dataset exhibits a near-linear relationship between the number of high-risk selections and peak infection reduction, suggesting that each additional intervention contributes roughly equally to mitigation. In contrast, the ABM network shows diminishing returns: even *λ* = 0.5 with *ϕ* = 1.0 yields ≈ 40% peak reduction, while increasing *λ* to 1.0 and 2.0 raises this to ≈ 55% and ≈ 70%, respectively. This pattern indicates diminishing returns as intervention intensity increases, suggesting that early targeting of the most connected individuals already captures much of the network’s epidemic potential. The DTU network lies between the two: low-sensitivity settings already yield ≈ 25% peak reduction, with gains rising more steeply with *λ* than in ABM but less so than in Office. These differences reflect structural features—Office is small and homogeneous, so interventions reduce transmission steadily, while the ABM is characterized by community structures, enabling early, high-impact targeting. The DTU network reflects real-world student mobility and shows bursty, layered contact patterns; its intermediate response profile likely reflects this mixture of structured and stochastic connectivity.

#### Efficiency Ratios: Normalized Gains per Intervention

Fig 4 shows both our efficiency ratios—peak-efficiency ℰ_peak_ and attack-rate efficiency ℰ_AR_)—as functions of the removal fraction *ϕ*, for all three datasets. In each panel, the dashed gray line indicates the “no-threshold” baseline in which every user is treated as high-risk at every intervention step.

**Fig 4.**
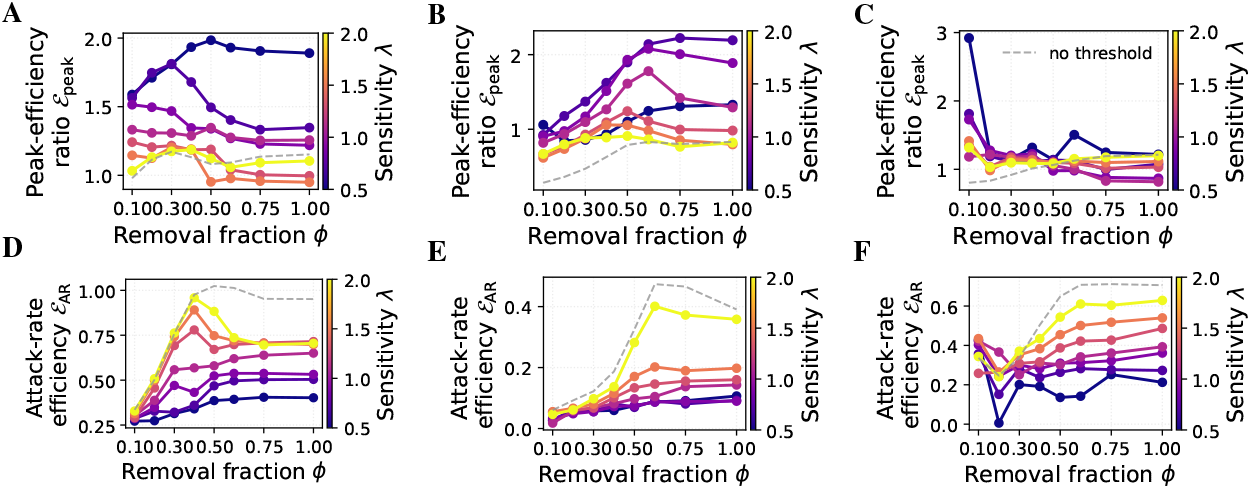
Efficiency ratios as a function of removal fraction *ϕ*. Results are shown for ABM (A, D), DTU (B, E), and Office (C, F). Top row: peak-efficiency ratio ℰ_peak_. Bottom row: attack-rate efficiency _AR_. Colors represent sensitivity *λ*, the dashed gray line indicates the no-threshold baseline, results are averaged over SIR runs.

In the ABM and DTU networks, the lowest sensitivity (*λ* = 0.5) consistently yields the highest peak-efficiency across the entire *ϕ* range, rising from about 1.5 (at *ϕ* = 0.1) to roughly 2.0 (at *ϕ* = 0.5), and then plateauing. For the ABM network, intermediate and higher *λ* values (e.g., *λ* = 1.0, 1.5, 2.0) start between ℰ_peak_ ≈ 1.0 and 1.5 at *ϕ* = 0.1, and tend to decrease steadily once *ϕ* exceeds 0.3–0.4. In contrast, for the DTU network, all *λ* values initially achieve ℰ_peak_ ≈ 1.0 at *ϕ* = 0.1, but increase with higher *ϕ* up to *ϕ* = 0.5, before stagnating or dropping slightly thereafter. This divergence reflects differences in network topology: in the ABM network, broader interventions decrease efficiency earlier, whereas in DTU, larger-scale interventions continue to yield proportional gains up to moderate *ϕ* levels. The Office network shows the same qualitative trend as ABM—low *λ* is slightly more efficient—but the effect is far less pronounced, reflecting its smaller, more homogeneous structure. Note the spurious spike to ℰ_peak_ ≈ 3 at *ϕ* = 0.1 in the Office case: because both the edge removal cost and the actual peak reduction are near zero when *ϕ* is very small, the ratio can become arbitrarily large despite negligible absolute impact. Across all datasets, the no-threshold baseline is the least peak-efficient, as it effectively mimics *λ* → ∞ and applies interventions to all individuals at every intervention step.

In contrast, the attack-rate efficiency *E*_AR_ (bottom row) exhibits nearly the opposite ordering. Here, higher sensitivities (*λ* ≥ 1.5) dominate: they achieve greater reductions in cumulative infections per edge removed, peaking around *ϕ ≈* 0.3 (ABM) or *ϕ ≈* 0.5 (DTU and Office). Again, the no-threshold baseline tracks closely with the *λ* = 2.0 line.

This divergence arises because our 10-day simulation window truncates the long-term dynamics: stronger, earlier interventions delay infections (flattening the curve) but, given more time, many of those infections would still occur once restrictions lift. Within a limited horizon, however, aggressive settings both slow transmission and keep cumulative case counts lower, inflating ℰ_AR_. Taken together, the two metrics illustrate a key trade-off: conservative, high-precision targeting (low *λ*) maximizes immediate peak reduction per contact removed, while more aggressive, lower-precision strategies (high *λ*) yield larger short-term reductions in total cases at the expense of higher social cost and potentially diminished long-term benefit.

The two metrics reflect different aspects of intervention impact rather than a strict trade-off. Low sensitivity (*λ* = 0.5) yields high ℰ_peak_ by selectively flattening the curve—delaying infections without greatly reducing total cases—so ℰ_AR_ remains low over our 10-day window. Stronger interventions (*λ* = 2.0) appear to boost ℰ_AR_ only because they suppress transmission long enough that cases don’t rebound within the limited horizon. In practice, unless interventions push the effective reproduction number below one (*R*_0_ < 1), delayed infections eventually occur, so ℰ_AR_ must be interpreted with care. By contrast, ℰ_peak_ directly captures the benefit of spreading cases over time—critical for managing peak healthcare demand.

### Risk and Intervention Distributions

Fig 5 summarizes three complementary views of NPCT’s behavior for *ϕ* = 0.25 on the ABM and DTU networks. Fig 5a shows the distribution of per-node risk scores 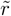 at the third intervention step for a fixed removal fraction *ϕ* = 0.25 of the ABM network. These counts are averaged over all SIR simulation runs and colored by sensitivity *λ*. For low *λ*, the histogram has a pronounced peak near 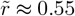. As *λ* increases, the entire distribution shifts leftward—higher sensitivity leads to a steeper drop in the threshold, causing more nodes to be classified as high risk and consequently broadening contact reduction, which in turn lowers the average population risk. In contrast, the risk distribution of the DTU network (Fig 5d) is more evenly spread across values between approximately 0.05 and 0.5, with a notably large mass near 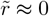. This reflects the higher heterogeneity of DTU’s contact structure, leading to a flatter, more uniform risk

**Fig 5.**
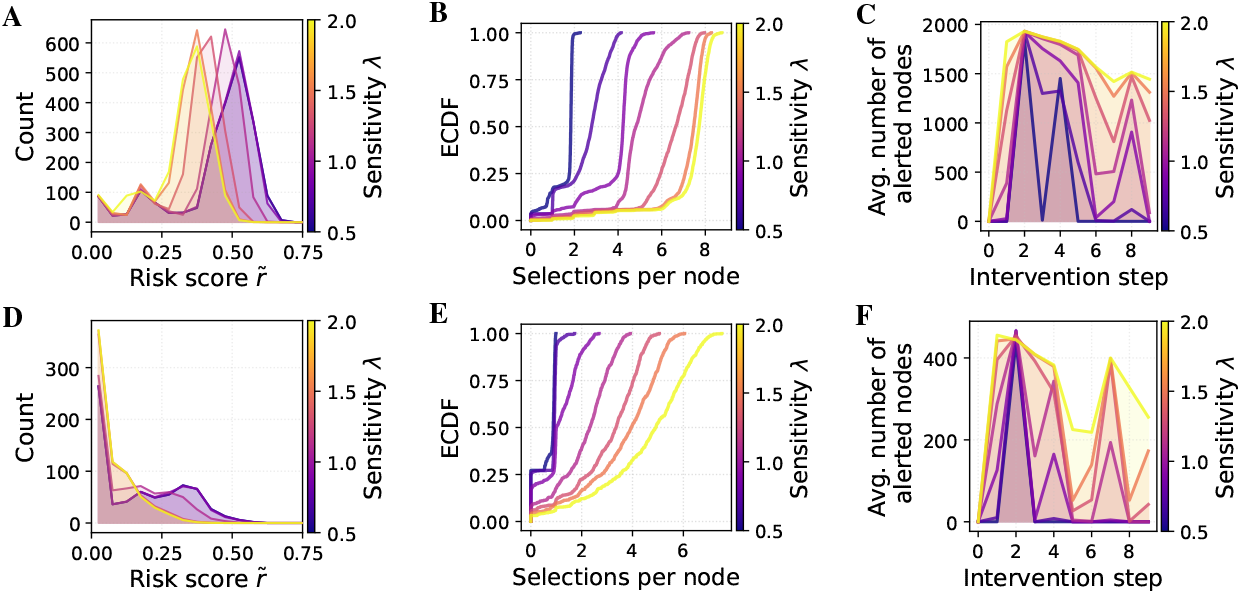
Risk and alert characteristics under NPCT. (A, D) Histogram of per-node risk scores 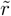 at intervention steps Δ*t* = 2 (*ϕ* = 0.25) and Δ*t* = 4 (*ϕ* = 0.50). (B, E) ECDF of total selections per node across ten interventions, for *ϕ* = 0.25 and *ϕ* = 0.50. (C, F) Number of alerted nodes at each step, for *ϕ* = 0.25 and *ϕ* = 0.50. Colors represent sensitivity *λ*, and results are averaged over SIR runs. Top row shows ABM, bottom row DTU results.

Fig 5b reports the empirical cumulative distribution function (ECDF) of the total number of alerts each node received over the 10-step horizon, for *ϕ* = 0.25. Each curve exhibits a flat portion followed by a sharp rise, indicating that a large group of nodes received the same number of alerts. As *λ* increases, the steep part of the curve shifts rightward: for low sensitivity (*λ* = 0.5), the majority of nodes are alerted just twice; for high sensitivity (*λ* = 2.0), they are alerted much more often. The presence of these steep transitions suggests relatively uniform intervention behavior within sensitivity levels—many nodes share the same alert count—while the shift in location reflects the overall increase in alert frequency with higher *λ*. The ECDF curves for the DTU network rise more gradually. Fig 5e shows that as *λ* increases, the curve flattens, indicating that the number of alerts is more heterogeneously distributed across nodes. This suggests that in the DTU network, different nodes are targeted with different frequencies depending on their local risk profile.

Fig 5c plots the average alert burden |ℋ (*T*_*j*_)| (number of nodes alerted) versus intervention step *j*. For low sensitivity values (around *λ* = 0.5), the curves exhibit sharp jumps at the second intervention, followed by strong relaxation in the next step, with the magnitude of these oscillations diminishing over time. At high sensitivity (*λ* = 2.0) there is a pronounced surge in alerted nodes at the first intervention, followed by only a gradual decrease in the average number of alerts over subsequent steps, reflecting a prolonged low threshold and sustained contact-reduction policy. In contrast, Fig 5f shows that the DTU network behaves comparably but with subtle differences. The sharp jump for *λ* = 0.5 at intervention step 2 appears only once and does not recur in subsequent steps, likely due to the network’s distinct temporal structure and the resulting epidemic trajectory. Nonetheless, similar to the ABM network, higher sensitivity values lead to a consistently elevated number of alerted nodes across interventions, with reduced relaxation compared to intermediate or low *λ* values.

Overall, we observe that the shape of the risk distribution 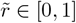 determines how the high-risk set responds as the threshold moves: if 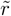 values are spread more evenly, as in the DTU network, the number of alerted nodes increases roughly linearly as the threshold declines. By contrast, the ABM network’s 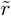 values cluster in a bell-shaped peak near 0.5, so many users share similar scores. The sensitivity parameter *λ* controls how finely the threshold adjusts: in a network with a tight cluster of 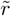 values, small changes in *λ* can cause large jumps in the high-risk set size. Consequently, for the ABM network, choosing *λ* requires extra care to avoid over- or under-alerting large cohorts of users simultaneously.

These results illustrate that increasing sensitivity *λ* not only shifts the population-level risk distribution to lower values and shifts the per-node alert-count curve but also sustains higher alert volumes over time, demonstrating how more a sensitive dynamic threshold yield earlier, wider, and more persistent interventions.

### Impact of user compliance

Fig 6 reports peak-infection reduction as a function of user-compliance level *c* ∈ *{*0.3, 0.6, 0.9, 1.0*}* at a fixed sensitivity *λ* = 0.7. Each panel shows the results for a network, with separate lines for each removal fraction *ϕ*.

**Fig 6.**
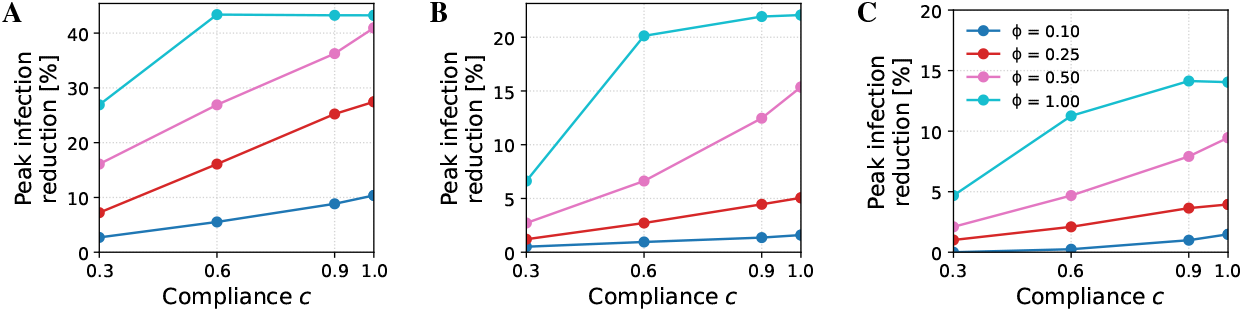
Peak infection reduction as a function of compliance *c*. Results are shown for ABM (A), DTU (B), and Office (C). Lines represent different removal fractions *ϕ*, results are averaged over SIR runs, and sensitivity is fixed at *λ* = 0.7.

Across all networks, the relationship is roughly linear: raising compliance from 0.3 to 1.0 steadily improves peak reduction. The gain is most pronounced when the prescribed removal fraction *ϕ* is large. For example, in ABM a jump from *c* = 0.3 to *c* = 0.6 at *ϕ* = 1.0 lowers the peak by nearly 15%, whereas the same compliance change at *ϕ* = 0.1 has only a marginal effect. This amplification effect is to be expected: when the baseline peak reduction is small (e.g. at low *ϕ*), scaling it by the compliance factor *c* produces only modest absolute gains, whereas interventions that remove more contacts offer greater headroom for improvements as compliance increases. As *c* → 1.0 the curves begin to plateau, especially at *ϕ* = 1.0: once shared contacts among high-risk users have been removed, additional adherence yields little extra benefit. Hence, perfect compliance maximizes impact, but even moderate adherence captures most of the flatten-the-curve benefit when contact reductions per user are substantial.

### Fairness

Fig 7 shows, for each of the three datasets (ABM, DTU, Office), the concentration curve of cumulative removal burden against cumulative risk share when the removal fraction is fixed at *ϕ* = 0.25. The horizontal axis represents the fraction of total risk contributed by the bottom *p*% of nodes, where risk is defined as the time-averaged, logarithmically normalized temporal degree 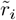 (Eq. 4). The vertical axis shows the fraction of total edges removed (the removal burden) carried by those same *p*% of nodes. The color of each curve encodes the sensitivity parameter *λ* (ranging from 0.5 to 2.0). Results for other removal fractions (*ϕ* = 0.10, 0.50, 1.00) appear in S4 Appendix.

**Fig 7.**
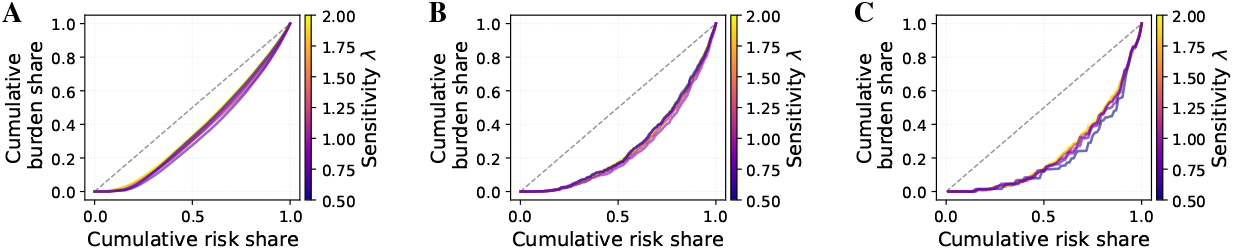
Concentration curves of cumulative removal burden vs. cumulative risk share for *ϕ* = 0.25. Results are shown for ABM (A), DTU (B), Office (C). The gray dashed line is the *y* = *x* reference for perfect equality. Colors represent sensitivity *λ*, results are averaged over SIR runs.

In all three panels the curves lie below the diagonal *y* = *x*, indicating that higher-risk nodes carry a disproportionately large share of the removal burden while lower-risk nodes incur proportionally little. The top 20% of the risk-ordered population (those beyond the 80th percentile) account for roughly 35% of all removals in ABM, about 40–45 % in DTU, and nearly 50% in Office, so almost half of the social cost is concentrated on the highest-risk quintile of users. Under a fairness notion that ties cost to epidemiological contribution, such a skew is desirable. Curves for different *λ* values are almost indistinguishable, demonstrating that sensitivity has negligible impact on burden distribution; likewise, varying *ϕ* produces minimal shift in this pattern (see S4 Appendix). This robustness follows from our dynamic threshold: once the highest-risk nodes have been intervened upon and their scores fall below the current cutoff, continued epidemic pressure drives the threshold downward—thereby adding intermediate-risk nodes in subsequent rounds rather than repeatedly targeting the same core.

### Long-horizon experiment: *ABM30*

To probe how NPCT behaves when the epidemic unfolds over a longer horizon, we re-ran our pipeline to generate a 30-day (**ABM30**) version of the ABM network introduced in the Datasets section. Because a longer horizon would otherwise saturate too quickly, we slowed the SIR process to *β* = 0.005, *γ* = 0.002.

Fig 8 confirms that the qualitative dynamics observed in the 10-day runs persist over the extended 30-day horizon. As the sensitivity parameter *λ* increases, the adaptive threshold responds more aggressively, triggering earlier and larger edge removals and yielding progressively stronger reductions in peak prevalence. The ordering of the prevalence curves and of the remaining-edge trajectories mirrors the shorter experiments, demonstrating that the feedback loop between epidemic pressure and threshold adaptation is robust to changes in epidemic timescale.

**Fig 8.**
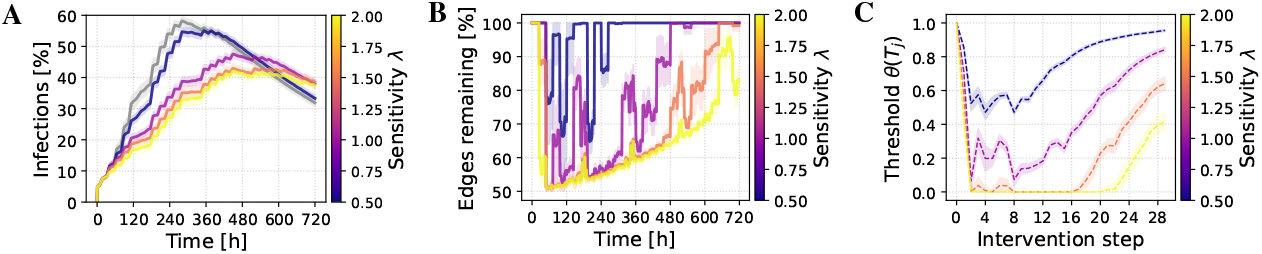
Progression of key quantities under NPCT interventions for ABM30 with *ϕ* = 0.25. (A) Infection prevalence over time (hours, x-axis; percentage infected, y-axis), with the gray curve representing the no-intervention baseline. (B) Percentage of edges remaining in the contact network (relative to baseline) over time (hours). (C) Evolution of the adaptive threshold *θ*(*T*_*j*_) across intervention steps. Colored curves represent sensitivity *λ* ∈ [0.5, 2], and results are averaged over SIR runs.

Fig 9 translates these dynamics into a quantitative cost-benefit landscape. In Fig 9b, we observe that the most efficient operating point—measured by the peak-efficiency ratio *E*_peak_—again arises for the most conservative setting *λ* = 0.5. Here *E*_peak_ climbs to roughly 2.0 at *ϕ ≈* 0.5 and then plateaus, matching the best values achieved on the day networks. For intermediate sensitivities, *λ* ∈ 1.0, 1.5, 2.0, the efficiency optimum shifts leftward to *ϕ* ≈ 0.25. These results reinforce the insight that conservative interventions yield the greatest benefit per contact removed, while more aggressive thresholds produce larger absolute peak reductions—exceeding 60% for *λ* = 2.0 (Fig 9a)—but with diminishing marginal returns. The attack-rate efficiency *E*_AR_ (Fig 9c) once again shows the opposite ranking, favoring higher *λ* values because they suppress the infection curve for a longer fraction of the temporal horizon.

**Fig 9.**
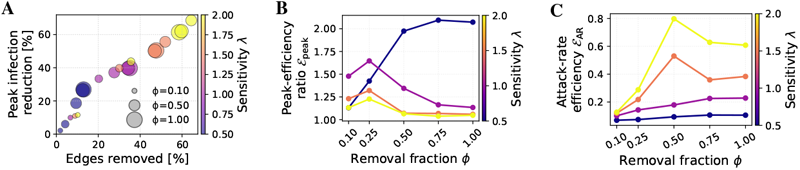
ABM30 efficiency landscape. (A) Bubble plot of peak reduction vs. edge removal, (B) peak efficiency and (C) attack-rate efficiency as functions of *ϕ*. Colors represent sensitivity *λ*, results are averaged over SIR runs.

Fig 10 displays the risk and intervention distributions of ABM30. Fig 10a shows the leftward shift of the risk histogram as *λ* grows, while Fig 10b illustrates how the cumulative alert count per user is displaced rightward. Fig 10c reveals a more nuanced alert-volume trajectory than in the 10-day runs: for *λ* = 0.5 and *λ* = 1.0 the number of alerted nodes oscillates smoothly. The longer horizon and reduced transmission rates dampen the extreme “all-on/none-on” oscillations reported in the Risk and Intervention Distributions section, allowing the threshold to explore intermediate regimes. This demonstrates that, with only two user-tunable parameters (*ϕ* and *λ*), NPCT can be calibrated to deliver a controllable spectrum of intervention intensities—ranging from highly selective to near-blanket suppression—without collapsing into pathological extremes.

**Fig 10.**
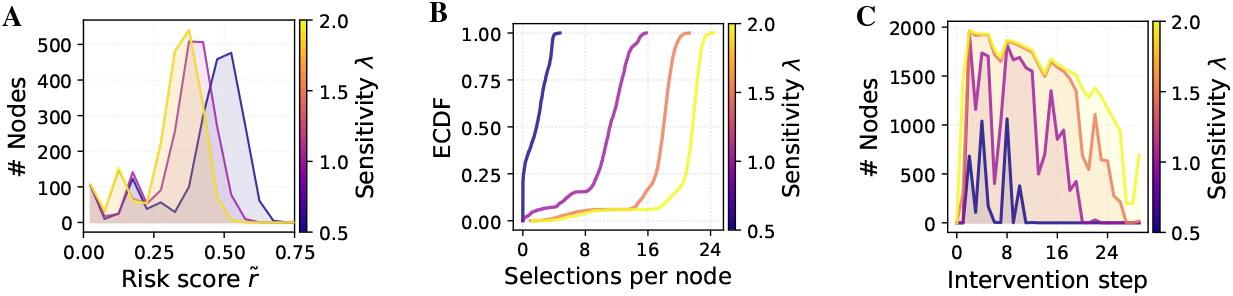
ABM30 risk and alert characteristics under NPCT for *ϕ* = 0.25. (A) Histogram of per-node risk scores 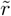 at intervention step Δ*t* = 2 (*ϕ* = 0.25). (B) ECDF of total selections per node across ten interventions for *ϕ* = 0.25. (C) Number of alerted nodes at each step for *ϕ* = 0.25. Colors represent sensitivity *λ*, and results are averaged over SIR runs.

### Limitations

While our study demonstrates the potential of Network-based Proactive Contact Tracing (NPCT), sparse data availability introduces constraints which should be considered when interpreting the results.

All empirical and synthetic benchmarks except the 30-day ABM30 network cover only ten days. Consequently, the core experiments in the Results section capture primarily short-term dynamics: how daily NPCT updates flatten the first infection peak. Long-term phenomena such as behavioral fatigue, decreasing immunity, seasonality, or multiple variant waves remain outside our empirical scope because no publicly available, high-resolution proximity datasets span multiple months. Given this temporal sparsity, we apply NPCT once every Δ*t* = 24 hours (see Simulation setup). A fixed, coarse cadence lets us trace how the adaptive threshold *θ*(*T*_*j*_) reacts to epidemic pressure and, crucially, keeps the intervention load comparable across datasets. Nevertheless, future studies should investigate different intervention windows.

As stated in the Intervention mechanism section, many individuals in real-world scenarios likely retain household or essential ties and drop specific contacts. This contact removal behavior and the resulting impact on NPCT efficiency should be further explored.

In the Intervention Efficiency section, we discussed that the peak-efficiency ratio *E*_peak_ inflates when both numerator (peak reduction) and denominator (edges removed) are near zero (e.g. Office network, *ϕ* = 0.1). Therefore, it is important to always consider absolute values alongside efficiency scores.

Despite these limitations, our results robustly demonstrate that a lightweight, privacy-preserving proactive layer can deliver substantial epidemiological benefits at modest social cost. As richer, longer-term datasets become available, the quantitative calibration of NPCT can be refined, but the core qualitative insight—that targeted, adaptive contact-suppression yields high efficiency—remains valid and promising for real-world deployment.

## Conclusion

Network-based Proactive Contact Tracing (NPCT) complements existing digital contact tracing deployments with an on-device early-warning layer that is at once privacy-preserving, lightweight and adaptive. Its risk engine relies solely on a user’s recent Bluetooth contact volume; its single, epidemic-driven threshold tightens or relaxes automatically in response to real-time acceleration and susceptible–infectious potential, removing the need for manual retuning. Extensive simulations on three ten-day networks and on a thirty-day extension show that NPCT can flatten the epidemic peak by roughly 40% while suppressing only about 20% of contacts, yielding a two-to-one return on social cost. The cost–benefit frontier is tunable: conservative sensitivities deliver the greatest benefit per edge removed, whereas aggressive settings push absolute peak reduction beyond 60% but with rapidly diminishing returns. The intervention’s overall behavior—both in how burden is distributed and in the volume of alerts—remains consistent across different sensitivity settings, removal strengths, compliance levels, and even over much longer time horizons. Across all networks, the sweet-spot removal strength fell in a narrow band: asking high-risk users to cut ≈ 25%–50% of their discretionary contacts captured most of NPCT’s peak-flattening benefit while avoiding the steep social costs that appear once *ϕ* approaches full quarantine. In practice, *λ* can remain a live dial—tuned by public-health dashboards.

These properties make NPCT a practical upgrade path for national exposure-notification apps: it preserves the cryptographic routines and decentralized data flows on which public trust depends, yet offers health authorities a transparent *λ*–*ϕ* dial to balance epidemic suppression against social disruption. Our analysis is based on a basic SIR model with a uniform population and employs random edge removal to represent contact reduction. In practice, incorporating vaccination status, age or risk groups, and more realistic patterns of how users actually modify their behavior could further refine both risk estimates and intervention outcomes. Taken together, our results demonstrate that a lightweight, privacy-preserving, network-aware early-warning layer can achieve substantial epidemiological gains at acceptable social cost—providing a clear roadmap for enhancing the next generation of digital contact-tracing tools.

## Data Availability

All contact-network datasets used in this study are publicly available. The Office network contains face-to-face proximity data collected in a French workplace and is available from the SocioPatterns repository at https://www.sociopatterns.org/datasets/office-proximity-network/. The DTU network represents the Bluetooth interaction layer of the Copenhagen Networks Study and can be accessed via Figshare at https://figshare.com/articles/dataset/The_Copenhagen_Networks_Study_interaction_data/7267433. The ABM network is available on Zenodo at https://doi.org/10.5281/zenodo.15076221. The ABM30 network is a 30-day extension of the ABM network and can be found at https://doi.org/10.5281/zenodo.15877149.

## Supporting information

**S1 Appendix. Nomenclature table**. Full list of model parameters and variables, including simulation, network, epidemic, and intervention-related quantities. See Table 2.

**Table 2.**
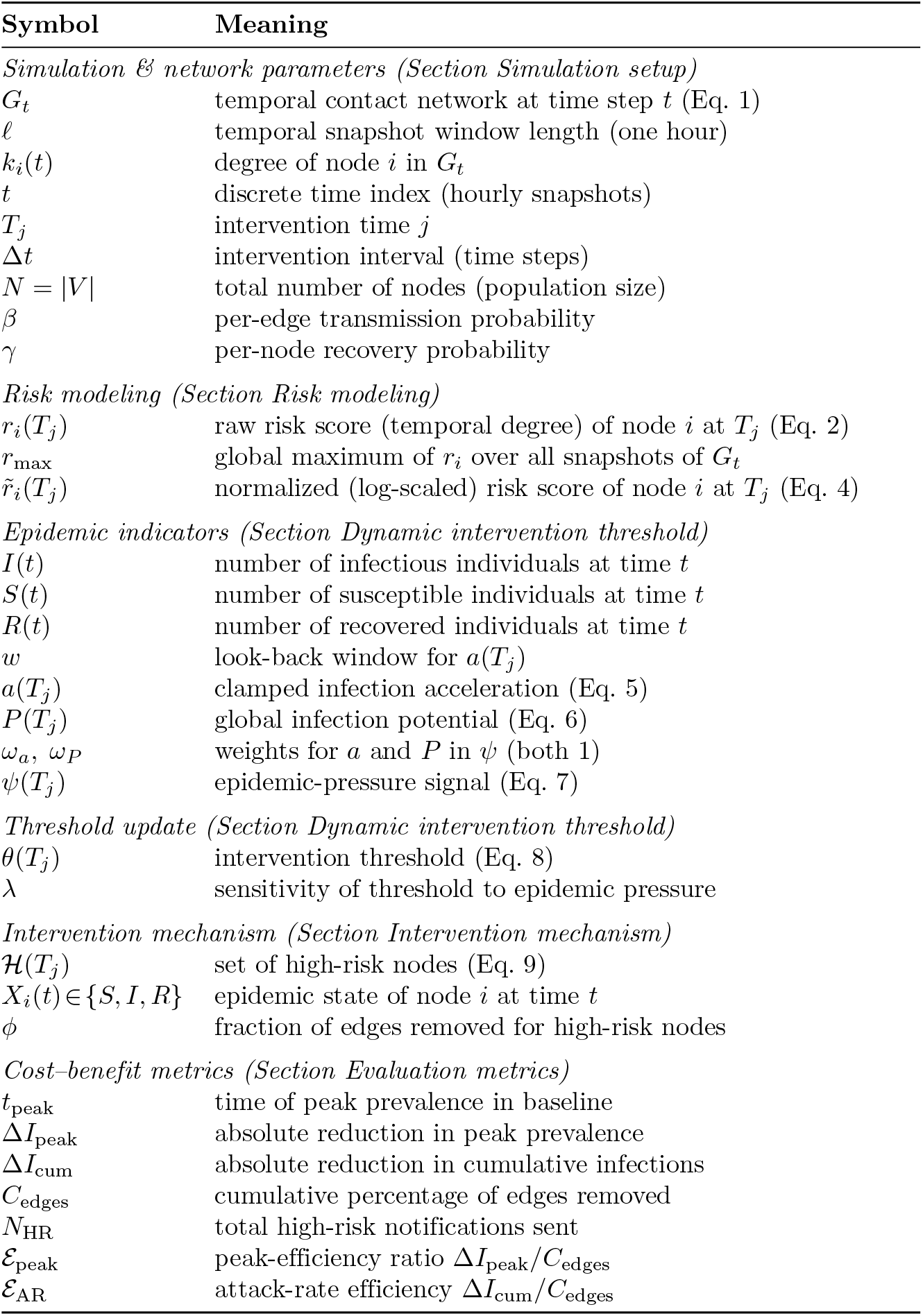
Complete list of parameters and variables.

**S2 Appendix. Beta–Gamma sweeps**. Heatmaps and infection curves across varying *β* and *γ* for three networks: ABM, DTU, and Office.

Fig 11 shows the exploration of the SIR parameter space for DTU, ABM and Office networks. Sweeping (*β, γ*)-pairs delimits the region of the plane that yields meaningful epidemic dynamics—avoiding trivial outcomes where the disease invariably dies out or infects the entire population—and guide our choice of transmission and recovery rates for the main experiments. We can observe a pronounced “phase-transition” band running diagonally across each heatmap: parameter combinations below this band correspond to outbreaks that fizzle out (final attack rates near 0 %), while those above it produce near-universal infection (final attack rates near 100 %).

**Fig 11.**
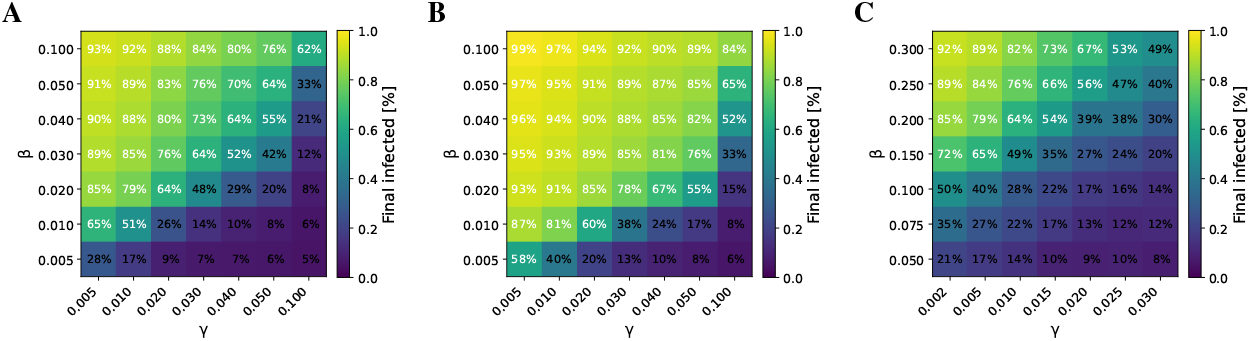
Heatmaps of the final proportion of infected individuals over the (*β, γ*) parameter space. Results are shown for ABM (A), DTU (B), and Office (C). For each subplot, the vertical axis is the transmission rate *β*, the horizontal axis is the recovery rate *γ*, and the color intensity indicates the percentage of the population ultimately infected.

As described in the SIR model section, we require each SIR simulation to exhibit a pronounced infection peak and at least 50 % prevalence at peak, so that the impact of our NPCT interventions is clearly visible. In Fig 12, we present additional (*β, γ*) pairs drawn from the band identified in Fig 11, from which we selected one representative combination per network. We note that the Office network displays larger variance in its epidemic curves—reflecting its highly dynamic contact patterns—while the DTU network sometimes shows a monotonic rise with no clear peak before the 10-day horizon. This underscores the importance of examining both peak timing and overall outbreak size, rather than final attack rate alone, when evaluating intervention efficacy.

**Fig 12.**
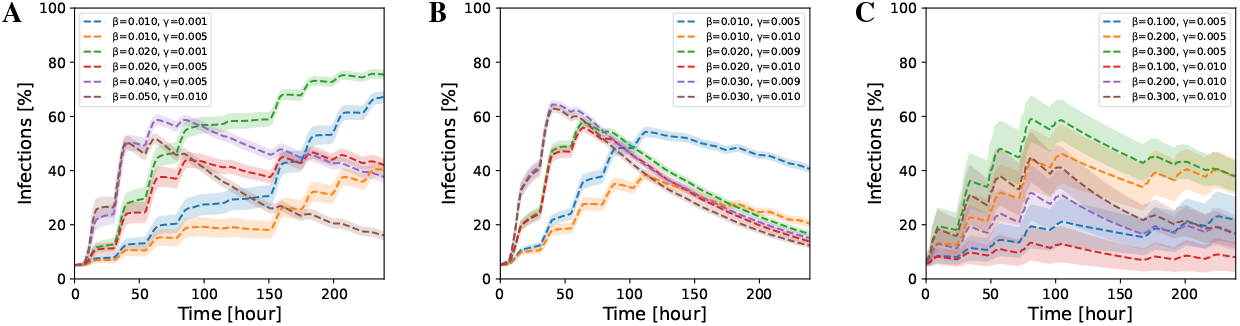
Epidemic curves from (*β, γ*) parameter sweeps. Results are shown for ABM (A), DTU (B), and Office (C). Each subplot shows the time evolution of the percentage of infected individuals under varying transmission rate *β* and recovery rate *γ*.

**S3 Appendix. Epidemic dynamics across networks**. Prevalence, edge count, and threshold tracking over time for the DTU and Office networks across different removal fractions *ϕ*.

For the DTU network (Fig. 13), the overall behavior closely mirrors what we observed on the ABM network. At low removal fractions (*ϕ* = 0.10, 0.25), the reduction in peak prevalence remains modest, but once *ϕ* = 0.50 is reached, a clear flattening of the infection curve appears for all sensitivity values *λ*. The adaptive threshold *θ*(*T*_*j*_) likewise follows the same characteristic trajectory seen in the ABM case, with increasingly pronounced oscillations at higher removal levels. These findings show that NPCT interventions are effective at slowing and attenuating outbreaks in a range of network types, but the specific temporal patterns and contact heterogeneity critically shape their impact—making it essential to tailor hyperparameters like the removal fraction *ϕ* and sensitivity *λ* to the network’s dynamic characteristics.

**Fig 13.**
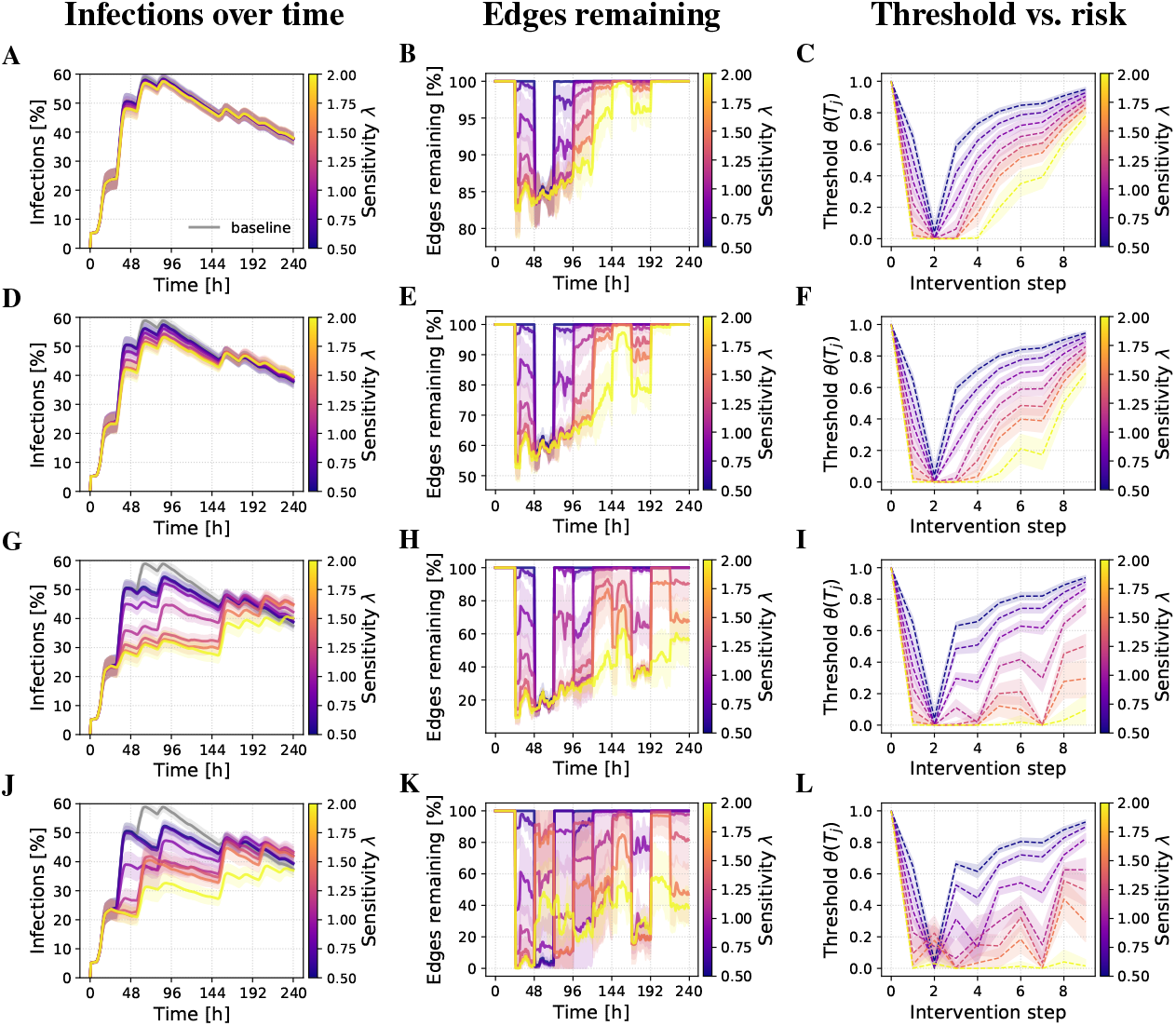
Progression of key quantities under NPCT interventions for DTU. (A, D, G, J) Infection prevalence over time (hours, x-axis; percentage infected, y-axis), with the gray curve representing the no-intervention baseline. (B, E, H, K) Percentage of edges remaining in the contact network (relative to baseline) over time (hours). (C, F, I, L) Evolution of the adaptive threshold *θ*(*T*_*j*_) across intervention steps. Colored curves represent sensitivity *λ* ∈ [0.5, 2], results are averaged over SIR runs. Each row corresponds to a removal fraction *ϕ* ∈ *{*0.10, 0.25, 0.50, 1.00*}* (top to bottom).

The Office network (Fig. 14) also exhibits a flattened epidemic curve under NPCT,but with an additional signature of its temporal structure: the middle panels tracing the fraction of remaining edges regularly jump back to 100% during off-hours, when no workplace contacts are recorded.

**Fig 14.**
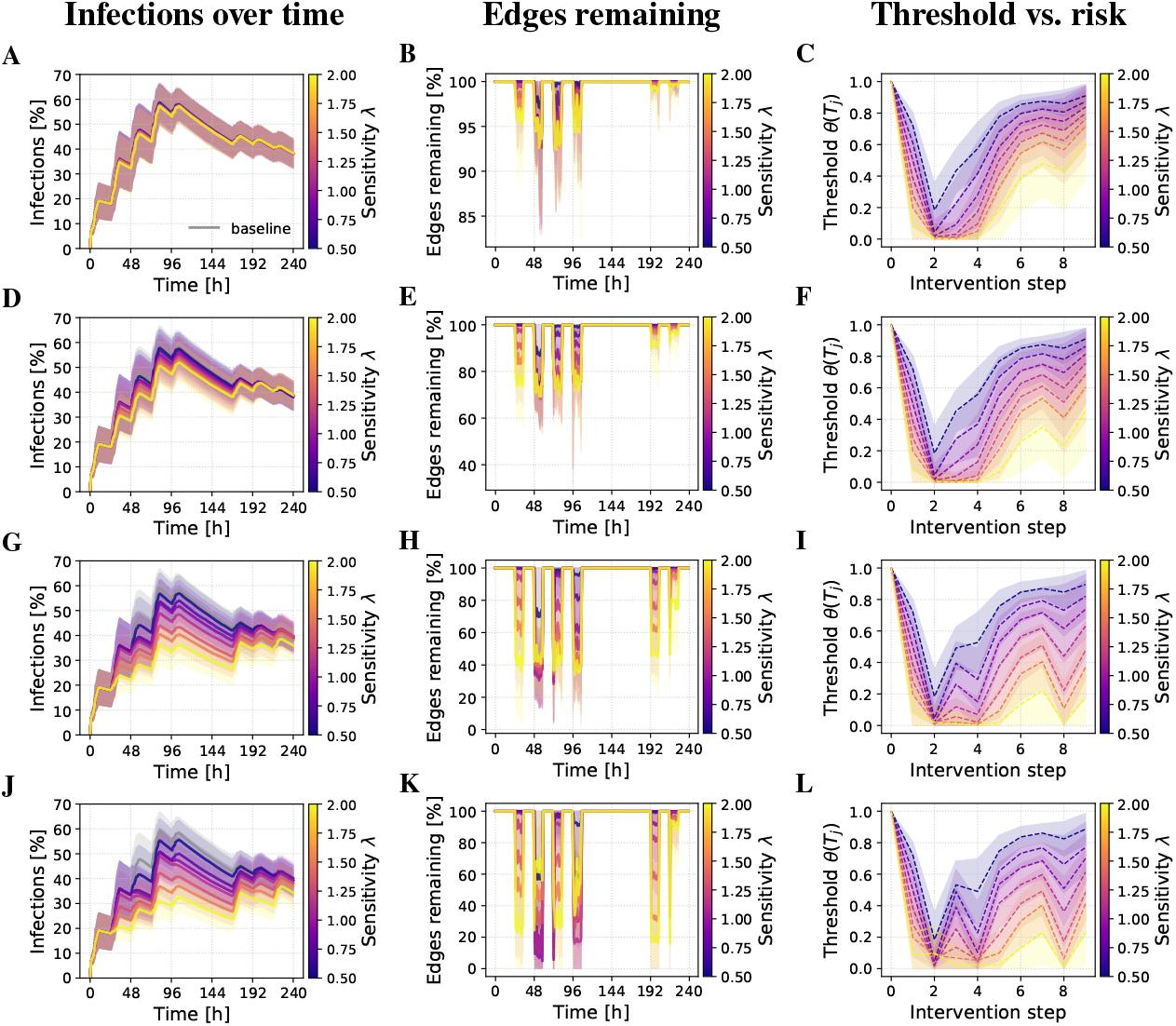
Progression of key quantities under NPCT interventions for Office. (A, D, G, J) Infection prevalence over time (hours, x-axis; percentage infected, y-axis), with the gray curve representing the no-intervention baseline. (B, E, H, K) Percentage of edges remaining in the contact network (relative to baseline) over time (hours). (C, F, I, L) Evolution of the adaptive threshold *θ*(*T*_*j*_) across intervention steps. Colored curves represent sensitivity *λ* ∈ [0.5, 2], results are averaged over SIR runs. Each row corresponds to a removal fraction *ϕ* ∈ *{*0.10, 0.25, 0.50, 1.00*}* (top to bottom).

**S4 Appendix. Burden distribution curves**. Concentration plots showing the burden of edge removals vs. risk score quantiles, stratified by *ϕ* and network type.

As shown in Fig 15, we have reproduced the concentration curves for three removal fractions (*ϕ* = 0.10, 0.50, 1.00) across the ABM, DTU, and Office networks. Consistent with the *ϕ* = 0.25 results, the curves for different sensitivity values *λ* remain almost indistinguishable, confirming that varying *λ* has minimal impact on how the removal burden is distributed by risk. A slight exception occurs at *ϕ* = 1.00 in the DTU and Office networks, where the *λ* = 2.0 curve shifts marginally closer to the equality line *y* = *x*, indicating a small move toward a more uniform workload. In the Office network panels, the curves also appear somewhat “stepped” rather than smooth, reflecting the smaller number of nodes: each individual removal constitutes a larger jump in the cumulative share, producing discrete increments. Even under full quarantine (*ϕ* = 1.00), however, the choice of sensitivity parameter does not substantially alter the overall fairness of the intervention.

**Fig 15.**
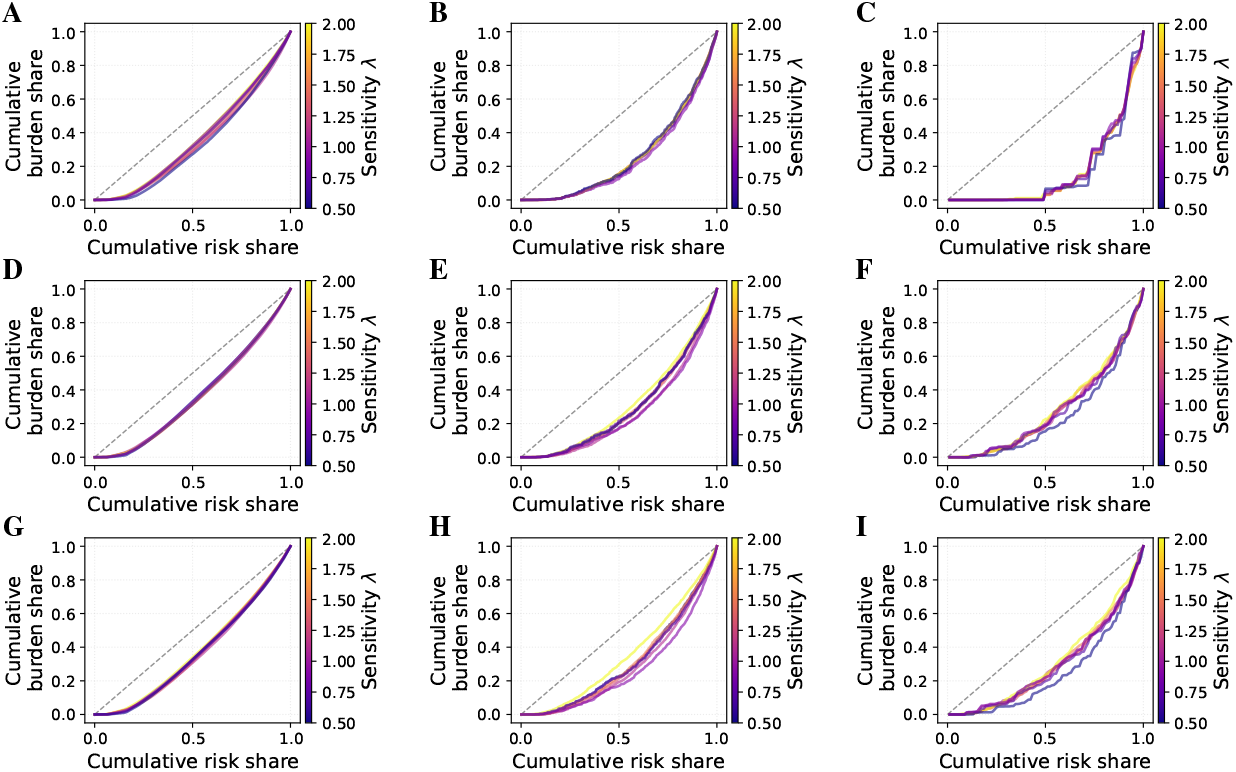
Concentration curves of cumulative removal burden vs. cumulative risk share for removal fractions *ϕ* = 0.10, 0.50, 1.00 across the ABM, DTU, and Office networks. The gray dashed line in each panel indicates perfect equality (*y* = *x*).

In Fig 16, we show for the Office network three diagnostics—risk distribution, selection ECDF, and alert-burden time series. We use the 5th intervention (Δ*t* = 4) and *ϕ* = 0.50. While the risk distribution for the Office network is skewed toward lower 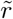 values compared to the DTU and ABM networks, the ECDF of node selections reveals a markedly broader spread of total selections per node, especially for higher removal fractions.

**Fig 16.**
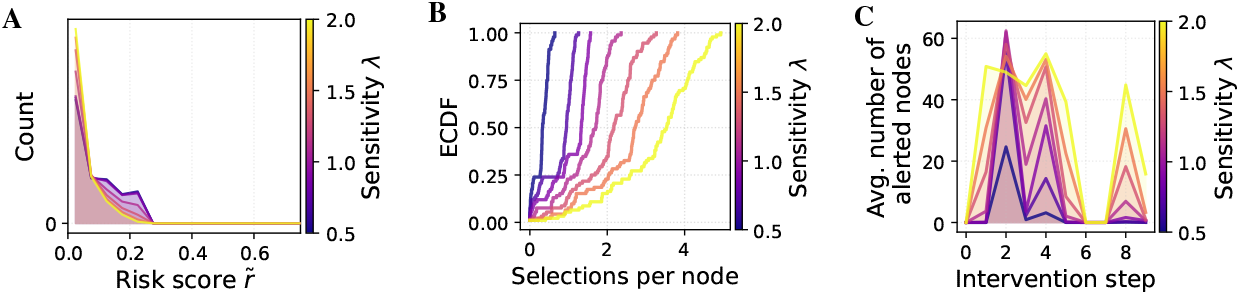
Risk and alert characteristics under NPCT for Office network. (A) Histogram of per-node risk scores 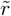 at intervention steps Δ*t* = 4 (*ϕ* = 0.5). (B) ECDF of total selections per node across ten interventions for *ϕ* = 0.5. (C) Number of alerted nodes at each step for *ϕ* = 0.50. Colors represent sensitivity *λ*, and results are averaged over SIR runs.

**S5 Appendix. Risk and intervention distribution diagnostics**. Risk histogram, ECDF of node selections, and alert burden over time for the Office network.

## References

1. Google LLC, Apple Inc. Exposure Notification API Bluetooth Specification v1.6; 2023. https://developers.google.com/android/exposure-notifications/exposure-notifications-api.

2. Ferretti L, Wymant C, Petrie J, Tsallis D, Kendall M, Ledda A, et al. Digital measurement of SARS-CoV-2 transmission risk from 7 million contacts. Nature. 2024;626(7997):145–150. doi:10.1038/s41586-023-06952-2.

3. Rodriguez P, Fernandez E, Rodon J, et al. A Population-Based Controlled Experiment Assessing the Epidemiological Impact of Digital Contact Tracing. Nature Communications. 2021;12:587. doi:10.1038/s41467-020-20817-6.

4. Segal CD, Lober WB, Revere D, Lorigan D, Karras BT, Baseman JG. Trading-off privacy and utility: the Washington State experience assessing the performance of a public health digital exposure notification system for coronavirus disease 2019. Journal of the American Medical Informatics Association. 2022;29(12):2050–2056. doi:10.1093/jamia/ocac178.

5. Bengio Y, Gupta P, Maharaj T, Rahaman N, Weiss M, Deleu T, et al. Predicting Infectiousness for Proactive Contact Tracing. arXiv preprint. 2020;.

6. Gupta P, Alsdurf H, Schmidt V, Qu M, Rahaman N, Bengio Y. Proactive Contact Tracing. PLOS Digital Health. 2023;2(3):e0000199. doi:10.1371/journal.pdig.0000199.

7. Hang CN, Tsai YZ, Yu PD, Chen J, Tan CW. Privacy-enhancing digital contact tracing with machine learning for pandemic response: a comprehensive review. Big Data and Cognitive Computing. 2023;7(2):108. doi:10.3390/bdcc7020108.

8. Marchetti S, Borin A, Conteduca FP, Ilardi G, Guzzetta G, Poletti P, et al. An epidemic model for SARS-CoV-2 with self-adaptive containment measures. PloS one. 2022;17(7):e0272009. doi:10.1371/journal.pone.0272009.

9. Yang H, Sürer Ü, Duque D, Morton DP, Singh B, Fox SJ, et al. Design of COVID-19 staged alert systems to ensure healthcare capacity with minimal closures. Nature communications. 2021;12(1):3767. doi:10.1038/s41467-021-23989-x.

10. Castillo-Laborde C, de Wolff T, Gajardo P, Lecaros R, Olivar-Tost G, Ramírez C H. Assessment of event-triggered policies of nonpharmaceutical interventions based on epidemiological indicators. Journal of Mathematical Biology. 2021;83:1–35. doi:10.1007/s00285-021-01669-0.

11. Di Lauro F, Kiss IZ, Rus D, Della Santina C. Covid-19 and flattening the curve: A feedback control perspective. IEEE Control Systems Letters. 2020;5(4):1435–1440. doi:10.1109/LCSYS.2020.3039322.

12. Kleinman RA, Merkel C. Digital contact tracing for COVID-19. CMAJ. 2020;192(24):E653–E656. doi:10.1503/cmaj.200922.

13. Ferretti L, Wymant C, Kendall M, Zhao L, Nurtay A, Abeler-Dörner L, et al. Quantifying SARS-CoV-2 transmission suggests epidemic control with digital contact tracing. science. 2020;368(6491):eabb6936. doi:10.1126/science.abb6936.

14. Endo A, Leclerc QJ, Knight GM, Medley GF, Atkins KE, Funk S, et al. Implication of backward contact tracing in the presence of overdispersed transmission in COVID-19 outbreaks. Wellcome open research. 2021;5:239. doi:10.12688/wellcomeopenres.16344.3.

15. Kojaku S, Hébert-Dufresne L, Mones E, Lehmann S, Ahn YY. The effectiveness of backward contact tracing in networks. Nature physics. 2021;17(5):652–658. doi:10.1038/s41567-021-01187-2.

16. Raymenants J, Geenen C, Thibaut J, Nelissen K, Gorissen S, Andre E. Empirical evidence on the efficiency of backward contact tracing in COVID-19. Nature Communications. 2022;13(1):4750. doi:10.1038/s41467-022-32531-6.

17. Schneider T, Dunbar OR, Wu J, Böttcher L, Burov D, Garbuno-Inigo A, et al. Epidemic management and control through risk-dependent individual contact interventions. PLOS Computational Biology. 2022;18(6):e1010171. doi:10.1371/journal.pcbi.1010171.

18. Briers M, Charalambides M, Holmes C. Risk scoring calculation for the current NHSx contact tracing app. arXiv preprint arXiv:200511057. 2020;.

19. Murphy K, Kumar A, Serghiou S. Risk score learning for COVID-19 contact tracing apps. In: Jung K, Yeung S, Sendak M, Sjoding M, Ranganath R, editors. Proceedings of the 6th Machine Learning for Healthcare Conference. vol. 149 of Proceedings of Machine Learning Research. PMLR; 2021. p. 373–390.

20. Rashidian M, Malek MR, Sadeghi-Niaraki A, Choi SM. Epidemic exposure risk assessment in digital contact tracing: A fuzzy logic approach. Digital Health. 2024;10:20552076241261929. doi:10.1177/20552076241261929.

21. Mason R. Covid: England facing weeks of ‘pingdemic’ disruption to services and food supply. The Guardian. 2021;.

22. Serafino M, Monteiro HS, Luo S, Reis SDS, Igual C, Neto ASL, et al. Superspreading k-Cores at the Center of COVID-19 Pandemic Persistence. PLoS Computational Biology. 2022;18(4):e1009865. doi:10.1371/journal.pcbi.1009865.

23. Sheikhahmadi A, Bahrami M, Saremi H. Minimizing Outbreak through Targeted Blocking for Disease Control: A Community-Based Approach Using Super-Spreader Node Identification. Scientific Reports. 2023;13:14217. doi:10.1038/s41598-023-41460-3.

24. Liang G, Cui X, Zhu P. An effective method for epidemic suppression by edge removing in complex network. Frontiers in Physics. 2023;11:1164847. doi:10.3389/fphy.2023.1164847.

25. CareEvolution. SAFER-COVID: A safe return to daily activities; 2020. https://careevolution.com.

26. Pandit JA, Radin JM, Quer G, Topol EJ. Smartphone apps in the COVID-19 pandemic. Nature Biotechnology. 2022;40(7):1013–1022. doi:s41587-022-01350-x.

27. Zhang X; MDPI. Decoding China’s COVID-19 health code apps: the legal challenges. Healthcare. 2022;10(8):1479. doi:10.3390/healthcare10081479.

28. Daugelaite T. China’s health code system shows the cost of controlling coronavirus. WIRED UK. 2020;.

29. Leitch J, Alexander KA, Sengupta S. Toward epidemic thresholds on temporal networks: a review and open questions. Applied Network Science. 2019;4:1–21. doi:10.1007/s41109-019-0230-4.

30. Keeling MJ, Eames KT. Networks and epidemic models. Journal of the royal society interface. 2005;2(4):295–307. doi:10.1098/rsif.2005.0051.

31. Pastor-Satorras R, Vespignani A. Epidemic spreading in scale-free networks. Physical review letters. 2001;86(14):3200. doi:10.1103/PhysRevLett.86.3200.

32. Heng K, Althaus CL. The approximately universal shapes of epidemic curves in the Susceptible-Exposed-Infectious-Recovered (SEIR) model. Scientific Reports. 2020;10(1):19365. doi:10.1038/s41598-020-76563-8.

33. Diallo D, Schoenfeld J, Schmieding R, Korf S, Kühn MJ, Hecking T. Integrating Human Mobility Models with Epidemic Modeling: A Framework for Generating Synthetic Temporal Contact Networks. Entropy. 2025;27(5):507. doi:10.3390/e27050507.

34. Diallo D, Schönfeld J, Blanken TF, Hecking T. Dynamic Contact Networks in Confined Spaces: Synthesizing Micro-Level Encounter Patterns Through Human Mobility Models from Real-World Data. Entropy. 2024;26(8):703. doi:10.3390/e26080703.

35. Sapiezynski P, Stopczynski A, Lassen DD, Lehmann S. Interaction data from the copenhagen networks study. Scientific Data. 2019;6(1):315. doi:10.1038/s41597-019-0325-x.

36. Génois M, Vestergaard CL, Fournet J, Panisson A, Bonmarin I, Barrat A. Data on face-to-face contacts in an office building suggest a low-cost vaccination strategy based on community linkers. Network Science. 2015;3(3):326–347. doi:10.1017/nws.2015.10.

37. Diallo D, Hecking T. Privacy-Preserving Vital Node Identification in Complex Networks: Evaluating Centrality Measures under Limited Network Information. In: Proceedings of the 2023 IEEE/ACM International Conference on Advances in Social Networks Analysis and Mining; 2023. p. 116–120.

38. Baunez C, Degoulet M, Luchini S, Pintus PA, Teschl M. Tracking the dynamics and allocating tests for COVID-19 in real-time: An acceleration index with an application to French age groups and départements. Plos one. 2021;16(6):e0252443. doi:10.1371/journal.pone.0252443.

39. Yu B, Chen X, Rich S, Mo Q, Yan H. Dynamics of the coronavirus disease 2019 (COVID-19) epidemic in Wuhan City, Hubei Province and China: a second derivative analysis of the cumulative daily diagnosed cases during the first 85 days. Global Health Journal. 2021;5(1):4–11. doi:10.1016/j.glohj.2021.02.001.

40. Utsunomiya YT, Utsunomiya ATH, Torrecilha RBP, Paulan SdC, Milanesi M, Garcia JF. Growth rate and acceleration analysis of the COVID-19 pandemic reveals the effect of public health measures in real time. Frontiers in Medicine. 2020;7:247. doi:10.3389/fmed.2020.00247.

